# Machine Learning Fairness in Predicting Underweight, Overweight and Adiposity Across Socioeconomic and Caste Group in India: Evidence from the Longitudinal Ageing Study in India

**DOI:** 10.1101/2025.07.06.25330978

**Authors:** John Tayu Lee, Sheng Hui Hsu, Vincent Cheng-Sheng Li, Kanya Anindya, Meng-Huan Chen, Charlotte Wang, Toby Kai-Bo Shen, Valerie Tzu-Ning Liu, Hsiao-Hui Chen, Rifat Atun

## Abstract

**Background:** Machine learning (ML) models are widely used to predict body mass index (BMI), yet their fairness across socioeconomic and caste groups remains uncertain, especially in countries with structure inequalities. This study evaluated the accuracy and fairness of ML models in predicting underweight, overweight, and central adiposity, examined the impact of socioeconomic and household factors, identified key predictive features, and assessed the effect of bias mitigation techniques on model performance.

**Methods:** This study analysed data from the nationally representative Longitudinal Ageing Study in India (LASI) with over 55,000 individuals aged 45 and older. We applied ML models (Random Forest, XGBoost, Gradient Boosting, LightGBM, DNN, DCN) alongside logistic regression. Model were trained (80%) and tested (20%), evaluated using AUROC, accuracy, sensitivity, specificity, and precision. Fairness assessment included subgroup analyses across socioeconomic and caste groups, equity-based fairness (e.g. Equalized Odds, Demographic Parity). Feature importance was examined using SHAP values. Bias mitigation techniques were applied at three stages: pre-processing (Disparate Impact Remover, Reweighting), in-processing (Exponential Gradient Reduction), and post-processing (Calibrated Equalized Odds, Reject Option Classification). Prediction density analysis assessed class separability across subgroup.

**Results:** Tree-based models—especially LightGBM and Gradient Boosting—along with Logistic Regression, consistently delivered the highest AUROC scores in predicting underweight, overweight, and high waist circumference outcomes (AUROC= 0.79-0.84). Incorporating socioeconomic and health-related variables gradually enhanced model performance; for example, the AUROC for underweight prediction increased from 0.74 to 0.78. However, our analysis revealed notable fairness issues: models performed worse for scheduled tribes and lower socioeconomic groups, as evidenced by reduced sensitivity and specificity in these subgroups. Feature importance analysis using SHAP values indicated that variables such as grip strength, gender, and residence were the key drivers of prediction differences; specifically, lower grip strength and rural residence were linked to underweight, whereas higher grip strength, urban residence, and female gender were associated with overweight and central adiposity.

Regarding bias mitigation, techniques like Reject Option Classification and Equalized Odds Postprocessing showed some potential for reducing subgroup disparities by aligning the performance of low- and high-performing groups. Nevertheless, these adjustments sometimes came with trade-offs, and other methods—such as Exponentiated Gradient Reduction and Adversarial Debiasing—resulted in substantial declines in overall performance. While approaches like Disparate Impact Remover, Reweighting, and the Stratified Subgroup Best Model produced only modest changes relative to the unmitigated model, our findings highlight persistent fairness challenges.

**Conclusions:** ML models can effectively predict obesity and adiposity risks in India, but addressing biases is critical for equitable application. There are needs to further refinement of fairness awareness ML approaches in public health, particularly in the context of India’s diverse population for more inclusive and effective policy decisions.

**AUTHOR SUMMARY:** India now faces the paradox of widespread under-nutrition alongside a rising tide of obesity among its older population. We asked whether state-of-the-art machine-learning models could accurately identify individuals at highest risk of under-weight, overweight–obesity, and central adiposity while treating all social groups equitably. Using nationally representative data on more than 55,000 adults aged 45 years and above, we compared gradient-boosted decision trees, random forests, logistic regression, and other approaches with conventional regression techniques.

Overall, the modern algorithms produced the strongest predictions. Yet a closer look revealed systematic shortfalls for scheduled tribes, scheduled castes, and the lowest income quintile—even when the models achieved excellent accuracy in the population as a whole. We then applied several well-established bias-mitigation strategies, such as re-weighting the training data and post-processing the decision thresholds. These interventions reduced the performance gap for disadvantaged groups, albeit at a modest cost to overall accuracy.

By combining careful fairness audits with Shapley-based interpretation of feature importance, we illuminate how socioeconomic and caste-related factors shape both nutritional risk and prediction error. Our findings underscore that fair, trustworthy decision support systems in public health must be designed explicitly with equity objectives, rather than assuming that technical excellence alone will guarantee just outcomes.

## INTRODUCTION

India, the most populous country in the world, faces significant health disparities across its diverse populations and geographic regions^1 2^. The prevalent issues of underweight, overweight, obesity, and adiposity are major contributors to these health inequalities and closed linked to country’s rapid demographic and epidemiological transitions^3–5^. These conditions range from malnutrition to overnutrition and mirror shifts in lifestyle, economic development, and societal changes throughout the nation^1 6 7^. This complex nutritional landscape imposes a significant burden of premature mortality and hinders progress toward multiple Sustainable Development Goals (SDGs), including those targeting good health and well-being (SDG 3), zero hunger (SDG 2), and reduced inequalities (SDG 10), as well as other interconnected goals such as quality education (SDG 4)^8^.

Early and accurate prediction of underweight, overweight, obesity, and adiposity is crucial for the effective implementation of public health interventions^9^. Extensive research has shown that the factors influencing these conditions vary significantly across India’s diverse populations^10–12^. This variability poses a challenge for traditional epidemiological methods, which often struggle to capture the complex interactions among sociodemographic, household, environmental, and lifestyle factors in detail. Machine learning approaches offer new solution with its advanced algorithms capable of adapting to and analyzing datasets with a wide range of interconnected variables. These methods not only improve our ability to characterize and predict body mass index (BMI)^13^ and anthropometric outcomes^14^ but do so with exceptional precision.

Moreover, in a variety of clinical domains, we can see that machine learning (ML) have led to remarkable improvements in predicting health outcomes—from diagnosis to risk stratification. For example, Rajkomar et al. (2018)^15^argued that ML systems have the potential to advance health equity if fairness is incorporated from design through deployment. Obermeyer et al. (2019)^16^demonstrated that widely used healthcare algorithms could exhibit substantial racial bias, leading to the under-identification of high-risk patients among marginalized groups. Despite these early insights, many subsequent studies have predominantly focused on overall predictive performance rather than rigorously assessing and mitigating bias across sensitive subgroups. A systematic review by Chen et al. (2024)^17^highlighted that although numerous methods exist to detect and reduce bias in machine learning models. It has been suggested that bias in machine learning models can arise from various sources, including the training data, the design of the algorithms, and the methods used to interpret data^15 17^. Such biases could potentially exacerbate existing disparities in health outcomes across different demographic groups, including variations across castes, and socioeconomic statuses^18–20^.

This study aims to evaluate the fairness in machine learning algorithm in predicting these BMI and central adiposity in India adult population. Specifically, the objectives are to (1) assess the accuracy and fairness of ML models across different socioeconomic and caste groups, (2) compare ML performance with traditional regression methods, (3) examine the impact of incorporating socioeconomic and household variables on model accuracy and fairness, (4) identify key predictive features influencing NML and central adiposity, (5) investigate the extent and source of bias in ML predictions across socioeconomic and caste groups, and (6) evaluate the effectiveness of various bias mitigation techniques in improving fairness without compromising model performance.

## RESULTS

### Descriptive Summary

The study included a total of 55,647 participants, of whom 46.67% were male and 53.33% were female. The participants’ ages ranged from 45 to 116 years, with a mean (SD) age of 59.4 (10.4) years. A demographic breakdown by caste indicated that 27.58% of participants belonged to the General category, 17.64% to the Scheduled Caste, 16.88% to the Scheduled Tribe, and 37.9% to Other Backward Classes. Another key variable, monthly per capita expenditure (MPCE), was categorized into five groups of approximately equal size: Lowest (20.28%), Lower Middle (20.33%), Middle (19.99%), Upper Middle (19.81%), and Highest (19.59%). In terms of health indicators, 18.5% (10,315 participants) were classified as underweight, 44.2% (24,570 participants) as overweight/obesity, and 45.9% (25,515 participants) exhibited a high waist circumference.

### Predictive Performance of Machine Learning Models

The predictive performance of various machine learning models for identifying underweight, overweight/obesity, and high waist circumference among the Indian population is summarized in *Supplementary Table 3*. *Figure 1* presents the ROC curves and corresponding AUROC values for these models. Notably, the ROC curve visualization and AUROC in the plot are based on the overall model applied to the test data without bootstrap resampling.

**Figure 1.**
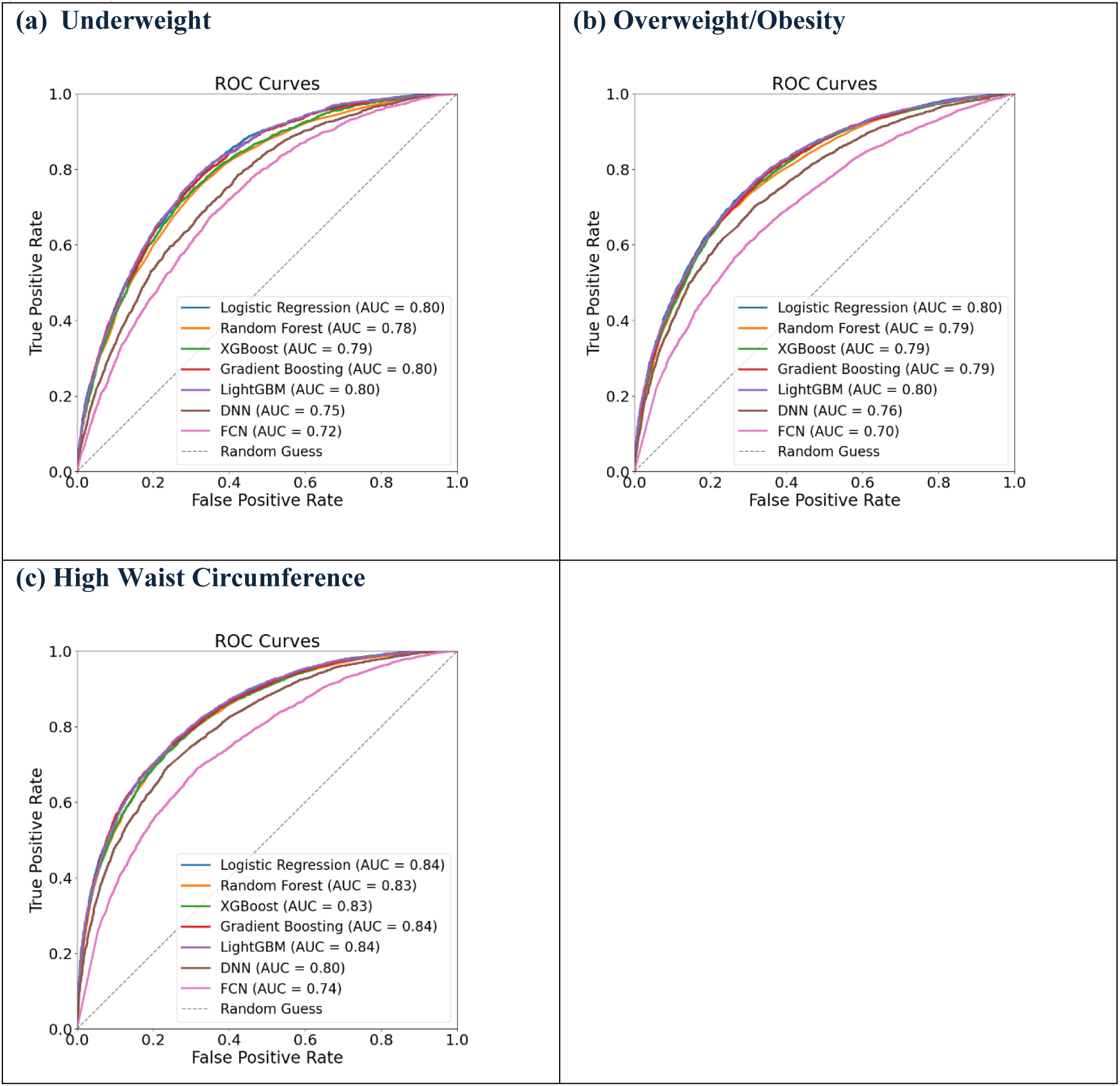
**Receiver Operating Characteristic (ROC) Curve with Area Under the Curve (AUROC) for Model Performance Evaluation**

#### Underweight Prediction

For underweight prediction, the AUROC values ranged from 0.72 to 0.80, with the highest performance observed in Logistic Regression, Gradient Boosting, and LightGBM, all achieving an AUROC of 0.80. While these models exhibited strong specificity, sensitivity remained consistently low across all groups. This discrepancy may be attributed to the imbalance in the data, as the prevalence of underweight cases was lower compared to other health outcomes. Additionally, models achieved higher accuracy for individuals with high MPCE compared to those with low MPCE; however, no notable difference was observed in AUROC.

#### Overweight/Obesity Prediction

For overweight/obesity prediction, the AUROC values ranged from 0.70 to 0.80, with LightGBM and Logistic Regression outperforming other models. However, the model’s performance varied across subgroups based on MPCE. In the lowest MPCE subgroup, LightGBM achieved a sensitivity of 0.37 (95% CI: 0.33–0.41) and a specificity of 0.92 (95% CI: 0.91–0.94), indicating limited ability to identify overweight/obesity cases despite strong non-case classification. Conversely, in the highest MPCE subgroup, sensitivity improved to 0.84 (95% CI: 0.82–0.86), but specificity dropped to 0.51 (95% CI: 0.47–0.54), highlighting a trade-off between case detection and non-case accuracy.

#### High Waist Circumference Prediction

For high waist circumference prediction, AUROC ranged from 0.74 to 0.84, with LightGBM, Gradient Boosting, and Logistic Regression again performing best. Similar to overweight and obesity prediction, the performance of LightGBM varied considerably across MPCE subgroups in terms of sensitivity and specificity, while accuracy remained relatively consistent. In the lowest MPCE subgroup, the model achieved a sensitivity of 0.52 (95% CI: 0.48–0.56) and a specificity of 0.89 (95% CI: 0.88–0.91). However, in the highest MPCE subgroup, sensitivity increased to 0.83 (95% CI: 0.80– 0.85), while specificity declined to 0.63 (95% CI: 0.60–0.66), reflecting a trade-off between improved case detection and reduced non-case classification accuracy.

#### Model Assessment Across Outcomes

Across all outcomes, Logistic Regression and LightGBM were identified as the most robust model overall, while neural network models consistently demonstrated lower performance. Due to its superior predictive capability across all outcomes and its ability to capture complex nonlinear relationships, LightGBM was selected as the primary model for further analysis.

### Prediction Density Analysis

Figure 2 shows the density plots of predicted probabilities for the positive (blue) and negative (orange) classes, stratified by caste and MPCE. Well-separated curves—often appearing as distinct double peaks—signal strong discrimination (high sensitivity and specificity), whereas overlapping curves indicate poorer performance.

**Figure 2.**
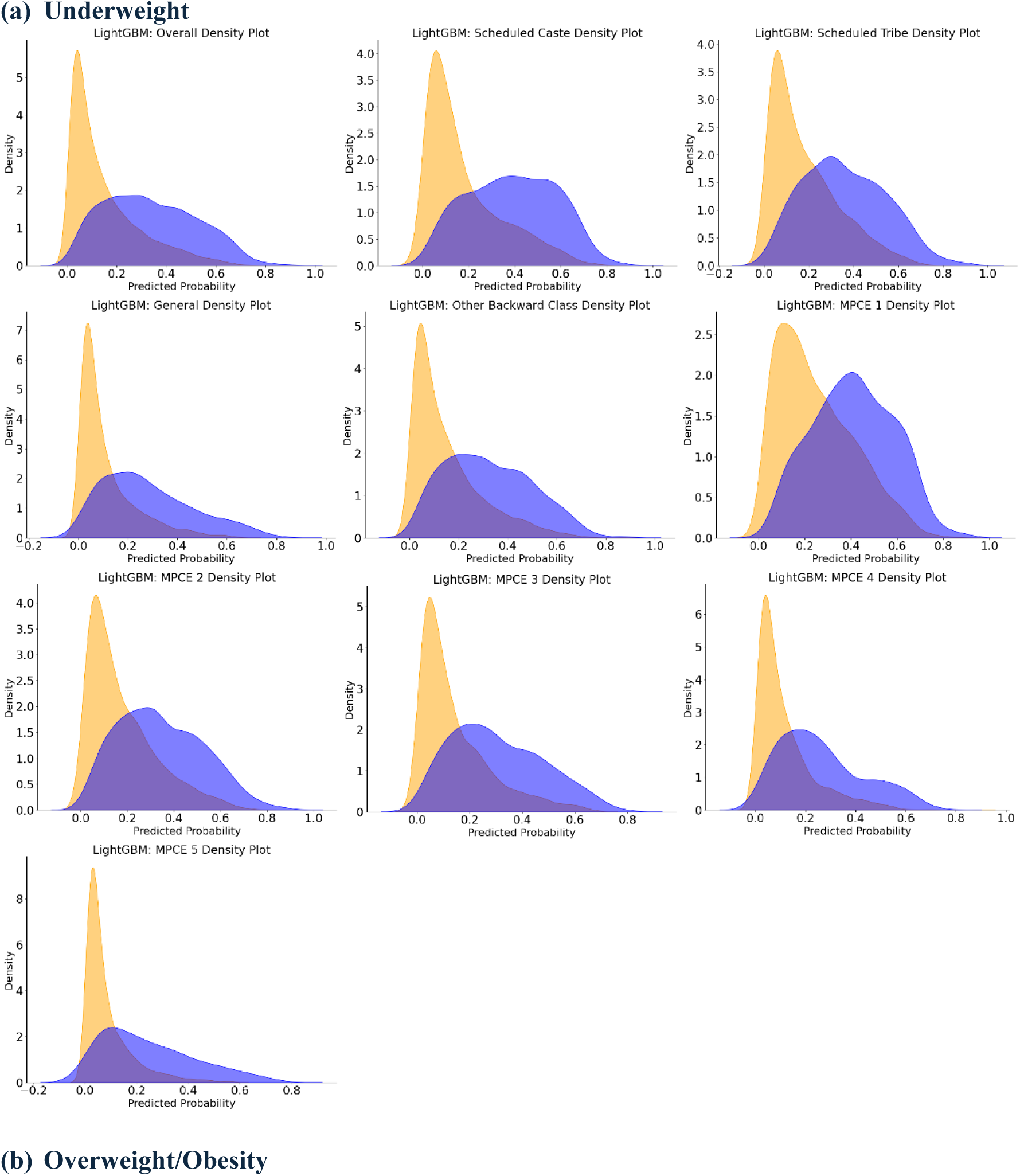
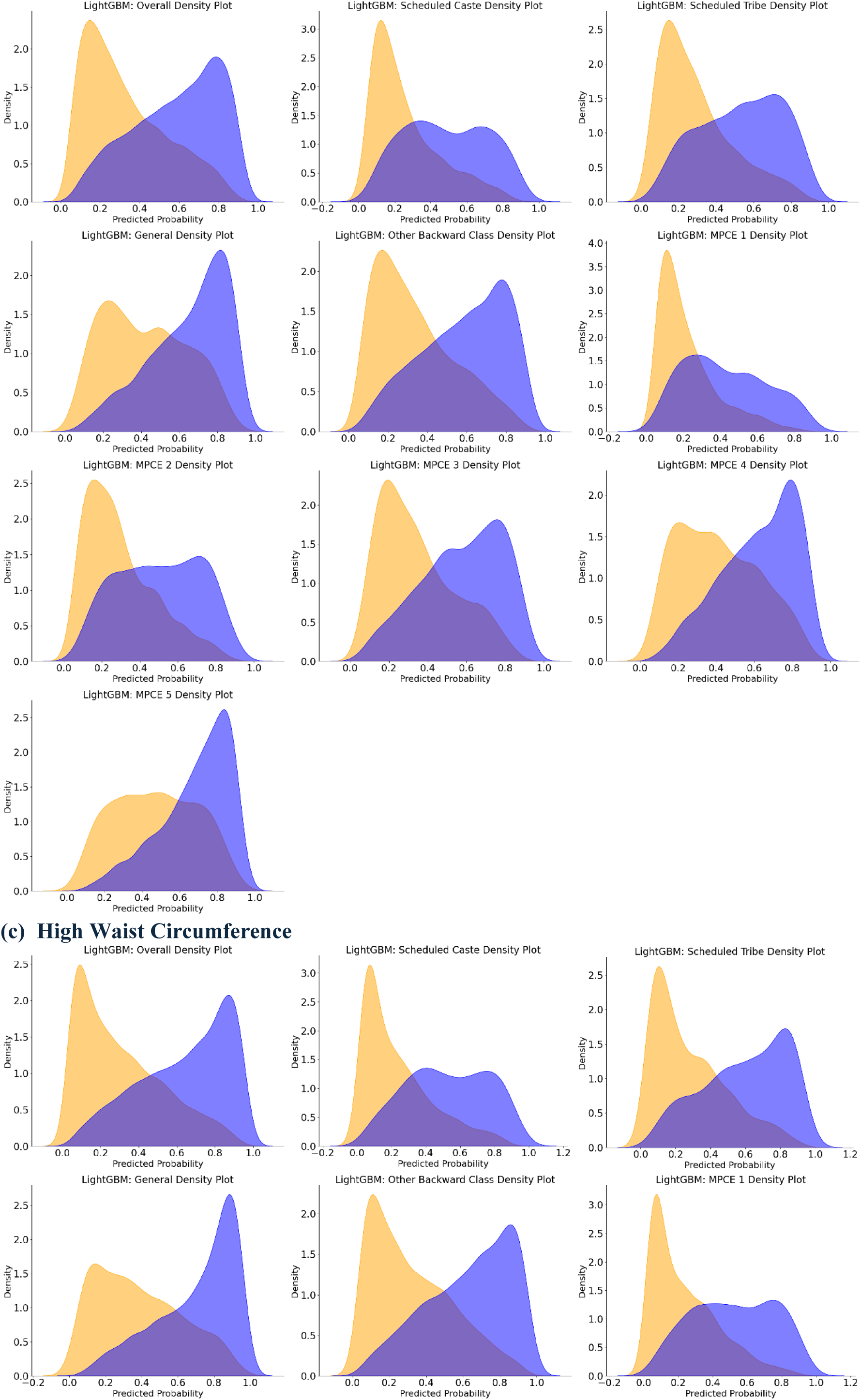
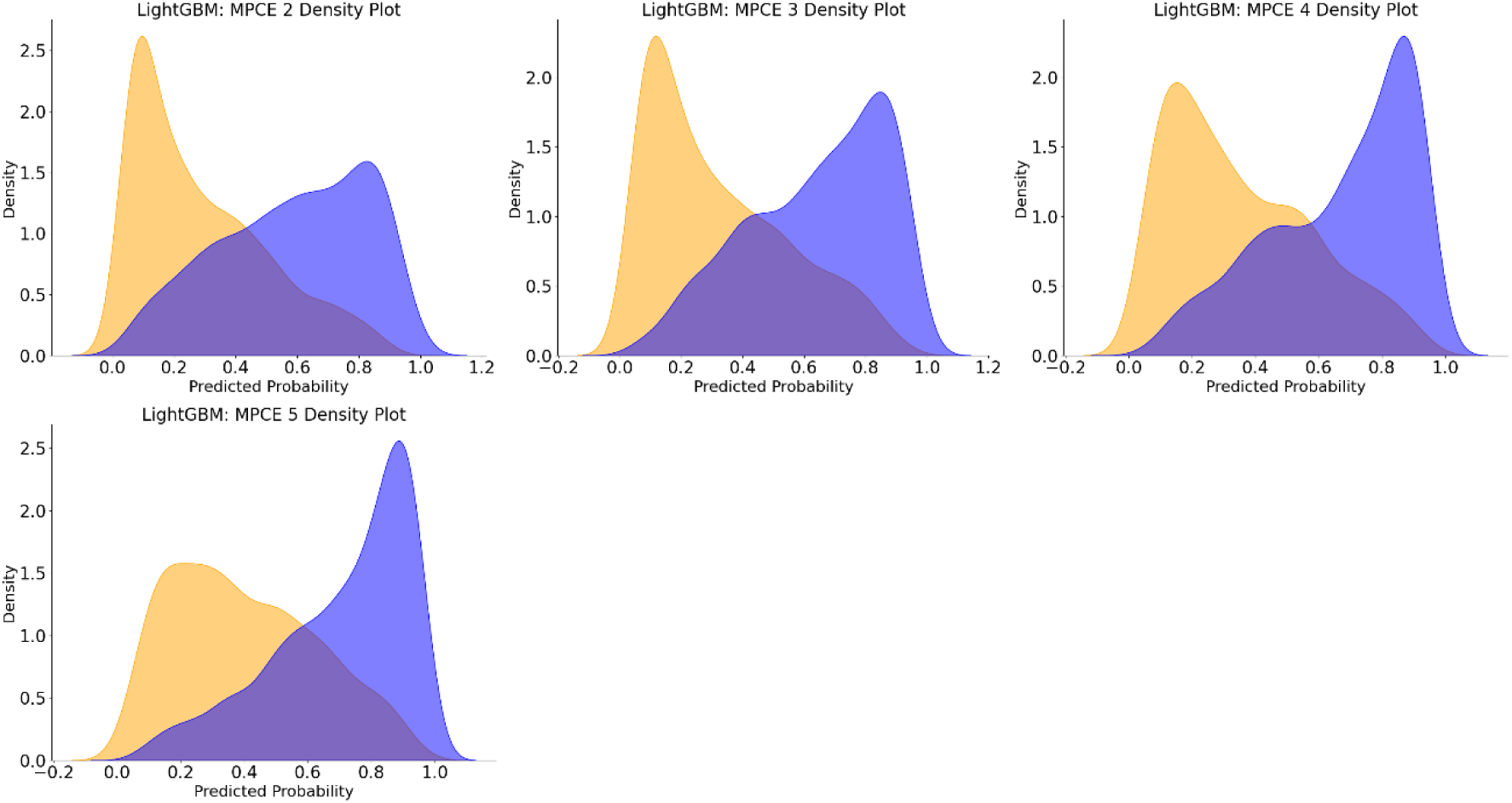
**Density Distribution of Model Predictions by Subgroups Stratified by Caste and Monthly Per Capita Expenditure (MPCE) Using LightGBM**

For underweight predictions, the positive and negative curves largely overlap across all subgroups, with only a slight left skew in the positive distribution, indicating weak discrimination. This is consistent with the lower sensitivity and specificity reported in Supplementary Table 2 for underweight predictions. In contrast, the overweight/obesity plots reveal more pronounced double peaks—especially in higher MPCE subgroups and among Other Backward Classes and General caste—though Scheduled Caste, Scheduled Tribe, and the lowest MPCE subgroup exhibit more overlap. A similar trend is observed for high waist circumference, where moderate to high MPCE subgroups show distinct double peaks, while the lowest MPCE subgroup displays considerable overlap; notably, the middle MPCE subgroup presents the clearest double peaks, suggesting the best balance between sensitivity and specificity.

Taken together, these density plots confirm that LightGBM discriminates overweight/obesity and high waist circumference more effectively than underweight, although performance varies by caste and socioeconomic status, with the poorest separation observed in the lowest MPCE and marginalized caste groups.

### Incorporating Socioeconomic and Health Features

Table 1 presents the performance metrics of the predictive models after incorporating both socioeconomic and health-related variables. For the underweight prediction model, the inclusion of these variables resulted in an increase in the AUROC from 0.74 to 0.75, and further to 0.78 after adding health-related variables. Similarly, for the overweight prediction, the AUROC increased from 0.75 to 0.76 with the addition of socioeconomic variables, and further to 0.78 after incorporating health-related variables. In the case of waist circumference, the AUROC improved from 0.80 to 0.81 with the inclusion of socioeconomic variables, and further to 0.82 with the addition of health-related data. These findings underscore the critical role of integrating socioeconomic and health-related factors to enhance the predictive performance of the models.

**Table 1.** Model Performance Metrics After Integrating Socioeconomic and Health-Related Variables Using LightGBM.

|  | AUROC | Accuracy | Sensitivity | Specificity | Precision |
| --- | --- | --- | --- | --- | --- |
| **Underweight** |  |  |  |  |  |
| Demographic | 0.7387 | 0.8100 | 0.0659 | 0.9856 | 0.5185 |
| + Socioeconomic | 0.7497 | 0.8144 | 0.0998 | 0.9830 | 0.5808 |
| + Health-related | 0.7756 | 0.8173 | 0.1482 | 0.9751 | 0.5844 |
| **Overweight/Obesity** |  |  |  |  |  |
| Demographic | 0.7493 | 0.6911 | 0.6013 | 0.7619 | 0.6659 |
| + Socioeconomic | 0.7612 | 0.7010 | 0.6203 | 0.7647 | 0.6753 |
| + Health-related | 0.7817 | 0.7140 | 0.6407 | 0.7719 | 0.6891 |
| **High Waist Circumference** |  |  |  |  |  |
| Demographic | 0.8050 | 0.7286 | 0.6534 | 0.7925 | 0.7284 |
| + Socioeconomic | 0.8136 | 0.7338 | 0.6720 | 0.7864 | 0.7282 |
| + Health-related | 0.8271 | 0.7487 | 0.6976 | 0.7922 | 0.7409 |

### Feature Importance Analysis

Figure 3 presents a summary plot of Shapley values for feature importance analysis. The most influential features were grip strength, gender, and residence, as indicated by the highest Shapley values. Additionally, hypertension, diabetes, and education level were also found to be significant contributors to the model predictions. These features played a crucial role in the overall performance of the predictive models.

**Figure 3.**
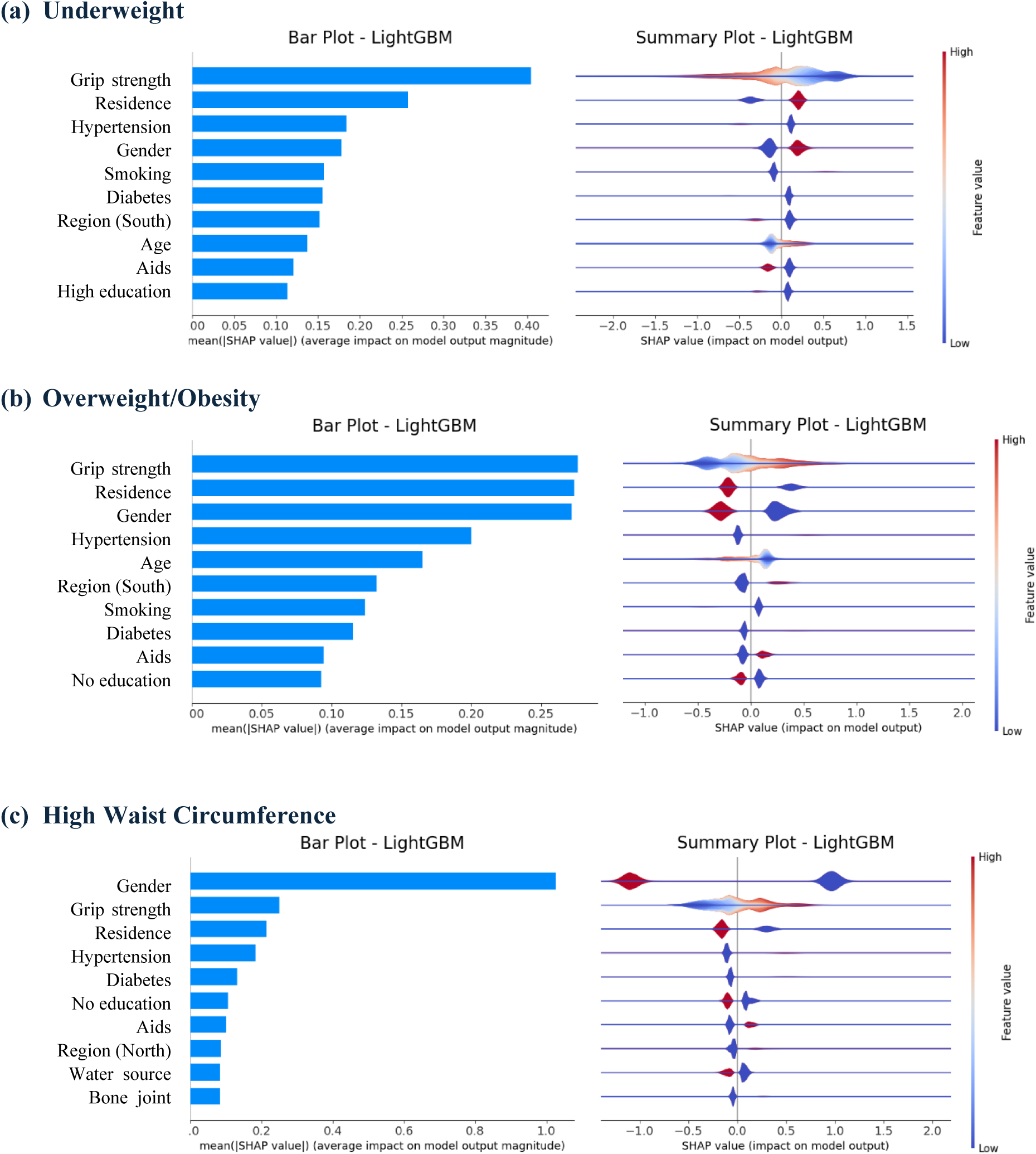
**Feature Importance Analysis and Summary Plot of Shapley Values for Model Interpretability**

Focusing on the top three features, we observed that grip strength had a negative impact on underweight individuals and a positive impact on overweight individuals and those with high waist circumference. Residence in rural areas was associated with a higher likelihood of underweight, while urban residents were more likely to be overweight or have a high waist circumference. Lastly, males were more likely to be underweight, while females were more likely to be overweight, with females also showing a higher likelihood of having a high waist circumference.

### Bias mitigation in Socioeconomic and Caste Subgroup

Figure 4, *Supplementary Table 4*, and *Supplementary Table 5* compared multiple bias mitigation strategies for the three target outcomes—underweight (Figure 4A1/A2), overweight/obesity (Figure 4B1/B2), and high waist circumference (Figure 4C1/C2)—by presenting box plots for three key performance metrics (AUROC, sensitivity, and specificity) across caste and MPCE subgroups. In these plots, the grey boxes represent the original best-performing model (LightGBM), while the colored boxes correspond to various bias mitigation techniques. Narrower boxes indicate smaller variability among subgroups (i.e., less disparity), and higher median values reflect stronger overall performance. Notably, Reject Option Classification and Equalized Odds Postprocessing (as seen in Figures 4A1 and 4A2) tend to “pull up” the low-performing subgroups and “pull down” the high-performing ones, which narrows the boxes and results in more uniform performance across caste and MPCE; however, this uniformity is sometimes achieved at the expense of lower median values, highlighting a trade-off between fairness and predictive power. In contrast, while Exponentiated Gradient Reduction and Adversarial Debiasing also reduce subgroup variability—especially in sensitivity—their median AUROC and sensitivity values drop substantially, indicating a more pronounced degradation in overall performance. Meanwhile, Disparate Impact Remover, Reweighting, and the Stratified Subgroup Best Model produce relatively modest changes, with box plots that remain close to the original grey distribution. Overall, Figure 4 underscores that although some methods can significantly narrow subgroup gaps—particularly for sensitivity and specificity— others introduce marked trade-offs in overall AUROC, demonstrating that different bias mitigation techniques vary widely in both their fairness benefits and their impact on predictive accuracy.

**Figure 4.**
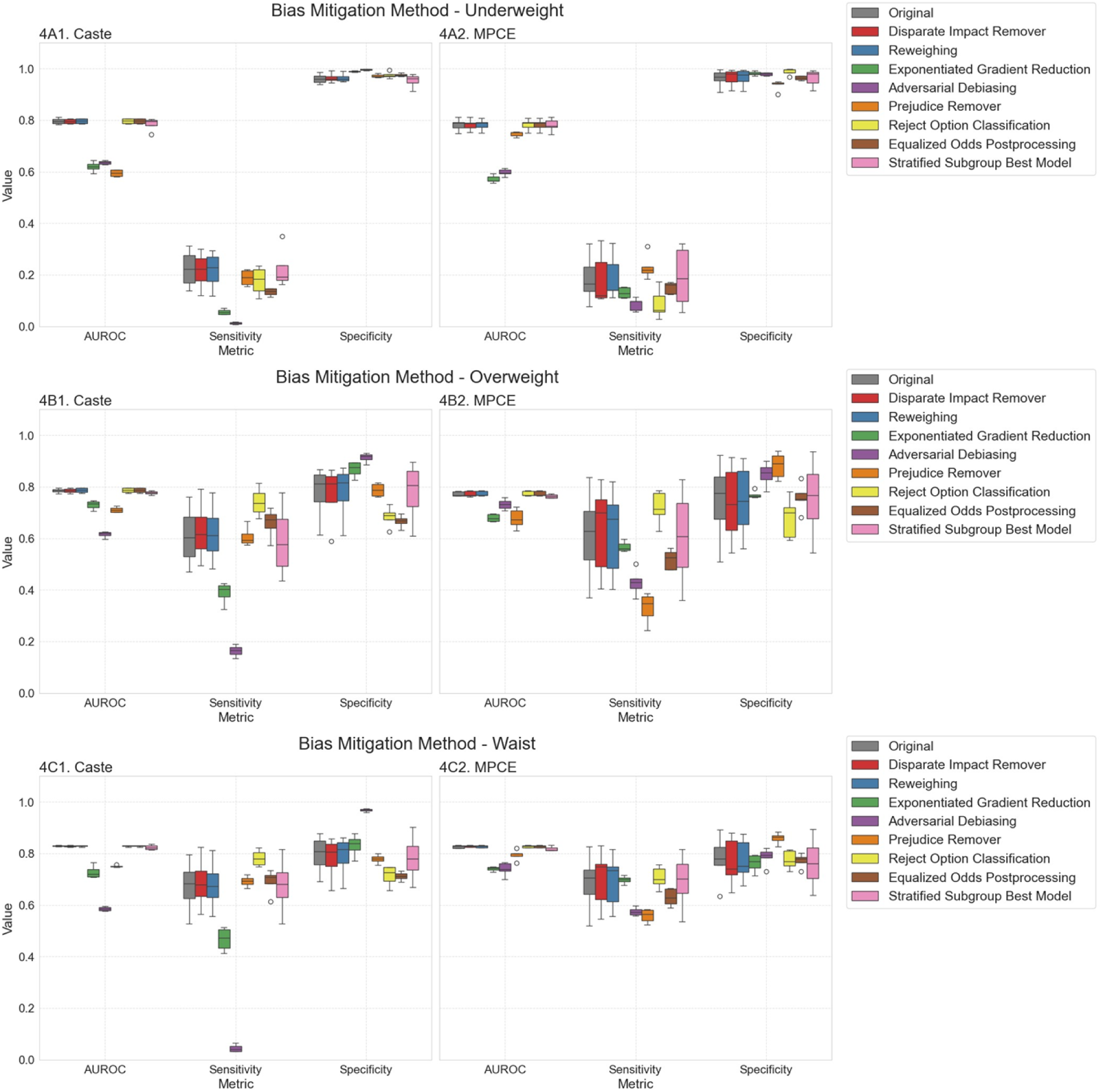
**Comparative Impact of Bias Mitigation Techniques on Subgroup Performance for Underweight, Overweight, and High Waist Circumference**

## DISCUSSION

This study is the first to evaluate the machine learning techniques for predicting underweight, overweight, obesity, and central adiposity within the Indian context, while also addressing biases that related to socioeconomic and demographic disparities. Our findings demonstrate that tree-based algorithms, particularly LightGBM and Gradient Boosting, outperform others in predicting anthropometric measures, including underweight, overweight, and waist measurements. Incorporating socioeconomic and health-related significantly enhanced model performance, highlighting their importance in improving model accuracy. Key predictors identified, such as grip strength, gender, residence, and hypertension, emphasize the role of socioeconomic and health variables in shaping anthropometric outcomes.

Despite overall strong predictive capability, performance varied across demographic groups, with marginalized populations, such as lower socioeconomic groups and scheduled tribes, experiencing lower accuracy. These findings underscore the need to address biases in ML models to ensure fair and equitable health predictions across diverse demographic groups.

Bias mitigation strategies, including Reject Option Remover and Equalized Odds Postprocessing, improved fairness, whereas Exponentiated Gradient Reduction and Adversarial Debiasing reduced disparities but at the cost of overall accuracy. Methods like Disparate Impact Remover, Reweighting, and the Stratified Subgroup Best Model yielded minimal improvements.

### Comparison with Existing Literature

Our study advances research by using machine learning to predict nutritional outcomes— underweight, overweight, obesity, and adiposity—in a large, nationally representative sample of older Indian adults. While previous studies focused on obesity prediction in specialized datasets, such as using deep learning on environmental factors^25^ or combining neuroimaging with metabolomics^26^, our work uniquely addresses nutritional risk in later life, where both under- and overnutrition coexist. Additionally, by incorporating culturally specific factors like socioeconomic status and caste, we provide a level of contextual nuance often overlooked in prior studies^27 28 29^.

Our approach is further distinguished by its rigorous evaluation and mitigation of algorithmic bias. Whereas prior studies using adversarial training have focused on reducing bias in models predicting outcomes like foregone care or cost-based risk among clinical populations^30^ and analyses of widely deployed risk prediction tools have revealed significant racial biases arising from label choice^16^ our work systematically implements a suite of debiasing techniques—including Disparate Impact Remover, Exponentiated Gradient Reduction, Re-sampling/Re-weighting, and Adversarial Training—complemented by stratified modeling. These measures help ensure that predictive accuracy is maintained equitably across marginalized subgroups (e.g., females, scheduled tribes, lower socioeconomic strata). In addition, although obesity risk prediction models developed in Bangladesh using relatively small datasets have shown high accuracy via logistic regression^31^ our study not only achieves competitive performance but does so while addressing fairness in a heterogeneous, underexplored older population. Collectively, our integrated framework of high predictive performance and robust fairness adjustment fills an important gap in the literature, offering actionable insights for equitable public health interventions in low- and middle-income settings.

### Policy Implications

Our findings highlight the potential of ML models to inform national health policies by predicting underweight, obesity, and adiposity with high accuracy. Algorithms like LightGBM show promise in predicting health outcomes such as underweight, obesity, and adiposity by incorporating socioeconomic and demographic variables. Key predictive features—grip strength, gender, residence, education, and chronic conditions—enable targeted, population-specific interventions. However, ML models often struggle struggle with performance disparities in marginalized groups, necessitating fairness-enhancing techniques to ensure equitable predictions. Addressing these gaps requires fairness-enhancing techniques, such as the Disparate Impact Remover, to ensure consistent model accuracy across diverse groups^15 17^.

This study has two main limitations. First, Our study did not use survey weights, as it is still unclear how to properly include them in machine learning studies^32^. Second, the use of self-reported data introduces potential biases, such as recall bias or social desirability bias, which could impact the accuracy of key variables. Future research should explore improved weighting methods and validate findings using additional datasets for broader generalizability.

## Conclusion

ML models offer significant promise in predicting obesity and related conditions in India, but addressing biases in predictions is crucial for equitable health outcomes. Our study underscores the need for fairness-driven ML approaches in public health, particularly in the context of India’s diverse population, providing actionable insights for more inclusive and effective policy decisions.

## MATERIALS and METHODS

### Data

This study uses a cross-sectional analysis of the Longitudinal Ageing Study in India (LASI) for the wave 2017-2018. LASI is a nationally representative survey of individuals aged 45 and above from all Indian states and union territories. The survey examines the health, social, and economic well-being of older adults. Harmonized with datasets like NHANES, ELSA, and CHARLS, LASI facilitates cross-population comparisons. Initially, the survey included 72,250 older adults and their spouses, enrolled through multistage probability sampling. After excluding 6,350 cases due to proxy interviews and participants younger than 45, the dataset was reduced to 64,867 individuals. Further data cleaning and processing to address incomplete records resulted in a final analytical sample of 55,647 participants.

The dataset provides detailed demographic, socioeconomic, and health-related information, including anthropometric measures such as Body Mass Index (BMI) and waist circumference (WC), collected through direct physical examinations. These data are critical for developing and validating machine learning models to identify obesity and related health risks. Data collection involved computer-assisted personal interviews, molecular biomarkers (e.g., dried blood spots), and non-molecular measures at the individual, household, and community levels. Variables used in this study are detailed in *Supplementary Table 2*.

### Measurements

The primary outcome measures include BMI and WC categories based on Asian standards for the Indian population: (1) BMI categories: underweight (<18.5 kg/m²), normal weight (18.5–22.9 kg/m²), and overweight/obesity (≥23 kg/m²). (2) WC cut-offs: Men: >90 cm; Women: >80 cm.

### Pre-processing of Independent Variables

The independent variables underwent systematic pre-processing to improve data quality and facilitate model interpretability. All pre-processing procedures, including handling missing data, variable encoding, standardization, and feature engineering, were documented and are accessible on GitHub (https://github.com/johntayuleeHEPI/LASI-BMI.git). More detail descriptions of all the variables can be found in *Supplementary Table 2*. After feature derivation, categorical variables were encoded using one-hot encoding, and rows containing missing values were excluded from the dataset.

### Machine Learning Algorithms

We employed multiple supervised machine learning (ML) models, including Random Forest, eXtreme Gradient Boosting (XGBoost), Gradient Boosting, Light Gradient Boosting Machine (LightGBM), Deep Neural Networks (DNN), and Fully Convolutional Networks (FCN). The dataset was divided into training (80%) and testing (20%) subsets, with the models trained on the training set and evaluated on the testing set. Features were standardized by calculating the z-score for each data point within the respective feature.

### Model Evaluation

The evaluation of model performance was conducted using the receiver operating characteristic (ROC) curve, a graphical representation of the trade-off between true positive rates (TPR) and false positive rates (FPR) across varying classification thresholds. The area under the ROC curve (AUROC) served as the primary metric to quantify the models’ discriminative ability, indicating their effectiveness in distinguishing between classes. The model with the highest AUROC was selected for further in-depth analysis.

To ensure a comprehensive evaluation, additional performance metrics, including accuracy, sensitivity, specificity, and precision, were computed. Notably, sensitivity and specificity were emphasized for their importance in identifying true positive and true negative cases relative to the ground truth. These metrics were further stratified and analyzed across subgroups to assess variations in model performance under different conditions.

All performance and fairness metrics were estimated using 1,000 bootstrap iterations to derive confidence intervals (CIs), ensuring robust and reliable estimates of the models’ capabilities and fairness across subgroups.

### Prediction Density Analysis

To evaluate the models’ discriminative power, prediction density analysis was conducted using density plots for positive and negative classes. These plots provide insights into the model’s sensitivity and specificity across subgroups. A left-skewed distribution for the positive class indicates high sensitivity, reflecting strong confidence in positive predictions, while a right-skewed distribution for the negative class signifies high specificity. In contrast, overlapping or flat distributions indicate poor class separation and limited predictive performance.

### Feature Importance Analysis

Feature importance was analysed using impurity-based feature importance and SHAP (SHapley Additive exPlanations) values. Impurity-based importance, common in tree-based algorithms, measures the reduction in impurity achieved by each feature. SHAP values, grounded in game theory, provide a detailed attribution of each feature’s contribution to individual predictions. These methods enhanced model interpretability by identifying key factors influencing predictions.

Additionally, features were categorized into three groups: demographic, socioeconomic, and health-related variables. Three models were constructed, each incorporating a different set of features: demographic-only, demographic + socioeconomic, and demographic + socioeconomic + health. This approach enabled the examination of how the inclusion of different feature categories impacted model performance and interpretability.

### Debias and fairness in Socioeconomic and Caste Subgroup

To address the biases identified during the validation phase, we modified the training data to ensure balanced representation across all groups. The bias mitigation techniques were applied in three distinct stages: pre-processing, in-processing, and post-processing.

In the pre-processing stage, we employed Disparate Impact Remover^21^, and Reweighting^22^ methods to adjust the dataset. In the in-processing stage, Exponential Gradient Reduction^23^ was applied to reduce bias during model training. For post-processing, we implemented fairness adjustments using Calibrated Equalized Odds Postprocessing^24^, Reject Option Classification, and Equalized Odds Postprocessing. These methods were implemented using IBM’s AI Fairness 360 (AIF360).

Additionally, we incorporated a stratified machine learning model to address potential arithmetic bias. The performance of this model was compared across various fairness adjustments to evaluate their overall impact on predictive accuracy. Notably, most bias mitigation approaches in our study relied on LightGBM following data pre-processing, with the exceptions of the Prejudice Remover—which defaults to logistic regression—and Adversarial Debiasing, which employs a deep learning framework. This integrated analysis enabled us to systematically assess the trade-offs between fairness and overall model performance.

### Patient and Public Involvement

Patients and/or the public were not involved in the design, conduct, reporting, or dissemination plans of this research.

### Ethics

The study was approved by the NTU Ethical Review Board (NTU REC-No.: 202407HS010). Ethical approval for LASI data collection was granted by the Indian Council of Medical Research and the Institutional Review Board (IRB) at IIPS, Mumbai. Informed consent was obtained at household and individual levels.

## AUTHOR CONTRIBUTIONS

JTL contributed the conception and design of this research, while SHH performed data extraction, model training, analytical methods, and results interpretation. KA assisted with data cleaning and extraction, and VCSL contributed to the literature review as well as revisions to the methods, results, and analysis code. MHC further supported the model training process and conducted the fairness analysis. Additionally, TKBS, VTL, and HHC collaboratively edited the manuscript, managed citations, and prepared the remaining sections, whereas RA and CW provided expert guidance on both the manuscript and the methodological design.

## Data Availability

The data that support the findings of this study are publicly available from https://github.com/johntayuleeHEPI/LASI-BMI.git

## SUPPLIMENTARY FILES

**APPENDIX TABLE**

**Supplementary Table 1.**
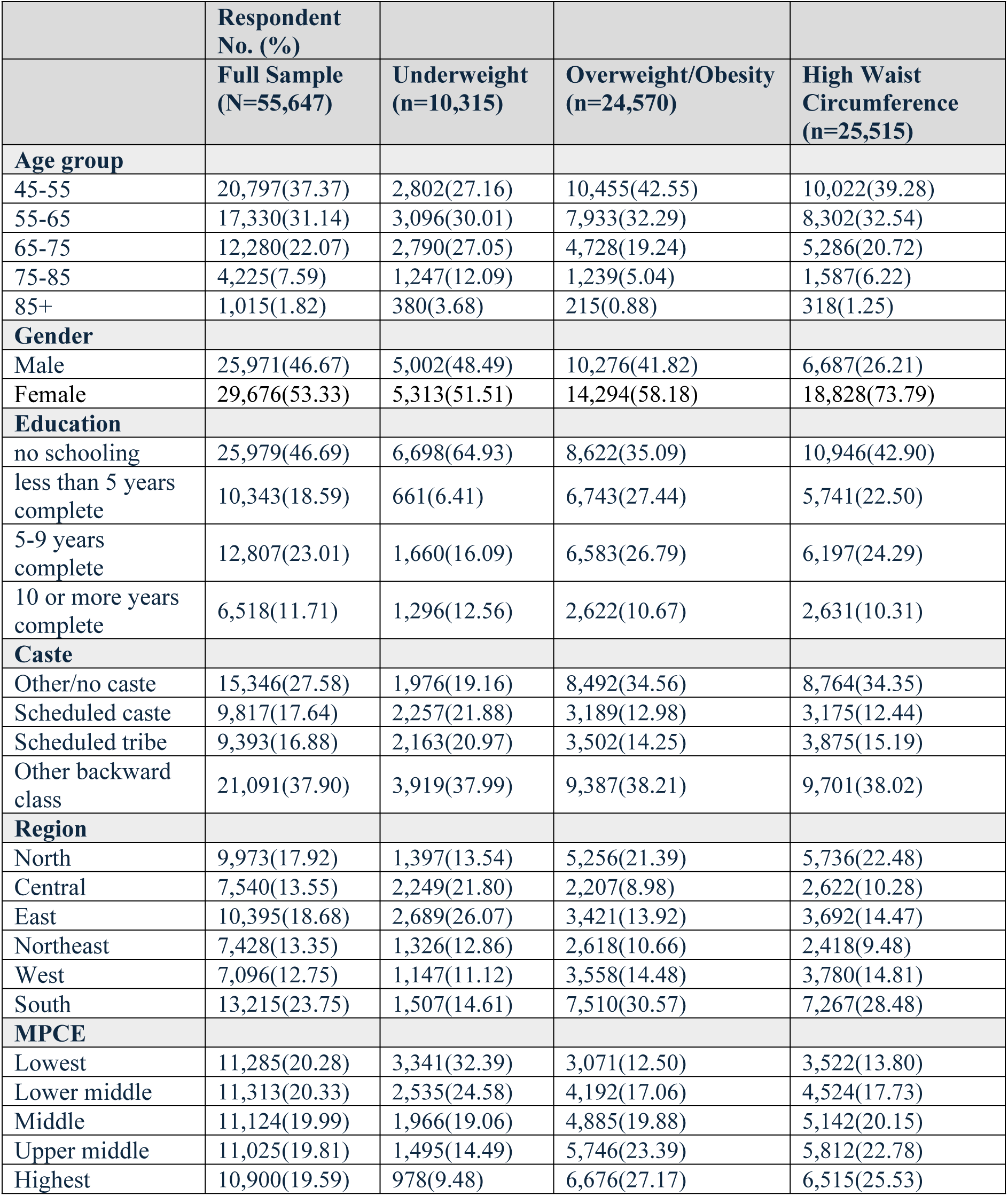
Descriptive Statistics of Key Health Outcomes: Prevalence of Underweight, Overweight, and High Waist Circumference Based on BMI and Waist Measurements.

|  | **Respondent**<br>**No. (%)** |  |  |  |
| --- | --- | --- | --- | --- |
|  | **Full Sample**<br>**(N=55,647)** | **Underweight**<br>**(n=10,315)** | **Overweight/Obesity**<br>**(n=24,570)** | **High Waist**<br>**Circumference**<br>**(n=25,515)** |
| **Age group** |  |  |  |  |
| 45-55 | 20,797(37.37) | 2,802(27.16) | 10,455(42.55) | 10,022(39.28) |
| 55-65 | 17,330(31.14) | 3,096(30.01) | 7,933(32.29) | 8,302(32.54) |
| 65-75 | 12,280(22.07) | 2,790(27.05) | 4,728(19.24) | 5,286(20.72) |
| 75-85 | 4,225(7.59) | 1,247(12.09) | 1,239(5.04) | 1,587(6.22) |
| 85+ | 1,015(1.82) | 380(3.68) | 215(0.88) | 318(1.25) |
| **Gender** |  |  |  |  |
| Male | 25,971(46.67) | 5,002(48.49) | 10,276(41.82) | 6,687(26.21) |
| Female | 29,676(53.33) | 5,313(51.51) | 14,294(58.18) | 18,828(73.79) |
| **Education** |  |  |  |  |
| no schooling | 25,979(46.69) | 6,698(64.93) | 8,622(35.09) | 10,946(42.90) |
| less than 5 years<br>complete | 10,343(18.59) | 661(6.41) | 6,743(27.44) | 5,741(22.50) |
| 5-9 years<br>complete | 12,807(23.01) | 1,660(16.09) | 6,583(26.79) | 6,197(24.29) |
| 10 or more years<br>complete | 6,518(11.71) | 1,296(12.56) | 2,622(10.67) | 2,631(10.31) |
| **Caste** |  |  |  |  |
| Other/no caste | 15,346(27.58) | 1,976(19.16) | 8,492(34.56) | 8,764(34.35) |
| Scheduled caste | 9,817(17.64) | 2,257(21.88) | 3,189(12.98) | 3,175(12.44) |
| Scheduled tribe | 9,393(16.88) | 2,163(20.97) | 3,502(14.25) | 3,875(15.19) |
| Other backward<br>class | 21,091(37.90) | 3,919(37.99) | 9,387(38.21) | 9,701(38.02) |
| **Region** |  |  |  |  |
| North | 9,973(17.92) | 1,397(13.54) | 5,256(21.39) | 5,736(22.48) |
| Central | 7,540(13.55) | 2,249(21.80) | 2,207(8.98) | 2,622(10.28) |
| East | 10,395(18.68) | 2,689(26.07) | 3,421(13.92) | 3,692(14.47) |
| Northeast | 7,428(13.35) | 1,326(12.86) | 2,618(10.66) | 2,418(9.48) |
| West | 7,096(12.75) | 1,147(11.12) | 3,558(14.48) | 3,780(14.81) |
| South | 13,215(23.75) | 1,507(14.61) | 7,510(30.57) | 7,267(28.48) |
| **MPCE** |  |  |  |  |
| Lowest | 11,285(20.28) | 3,341(32.39) | 3,071(12.50) | 3,522(13.80) |
| Lower middle | 11,313(20.33) | 2,535(24.58) | 4,192(17.06) | 4,524(17.73) |
| Middle | 11,124(19.99) | 1,966(19.06) | 4,885(19.88) | 5,142(20.15) |
| Upper middle | 11,025(19.81) | 1,495(14.49) | 5,746(23.39) | 5,812(22.78) |
| Highest | 10,900(19.59) | 978(9.48) | 6,676(27.17) | 6,515(25.53) |

**Supplementary Table 2.** Definitions and Descriptions of Variables Used in the Analysis.

| Variable Name | Type | Definition/Categories |
| --- | --- | --- |
| **Demographic Variables** |  |  |
| Age | num | Continuous age measured in years. |
| Gender | binary | Categories: Male; Female. |
| Education | category | Categories: No schooling; Less than 5 years complete; 5–9 years complete; 10 or more years complete. |
| Migration status | binary | Categories: Yes (migrated); No (non-migrated). |
| State | category | 35 states in India. |
| Region | category | Categories: North; Central; East; Northeast; West; South. |
| Residence | binary | Categories: Rural; Urban. |
| Religion | category | Categories: Hindu; Muslim; Others. |
| Marital status | binary | Categories: Yes (married); No (unmarried/divorced/widowed). |
| Living alone | binary | Categories: Yes (living alone); No (living with others). |
| **Socioeconomic Variables** |  |  |
| Economic status (MPCE quintile) | category | Categories: Lowest; Lower middle; Middle; Upper middle; Highest. |
| Working status | category | Categories: Currently working; Never worked; Have worked in the past. |
| Type of main job | category | Categories: Agricultural/Fishery/Labor; White-collar job; Others; No current work. |
| Monthly pension | num | Continuous monthly pension amount. |
| Retired officially | binary | Categories: Yes (officially retired); No (not retired). |
| Pension received | binary | Categories: Yes (receiving pension); No (not receiving pension). |
| Caste | category | Categories: General; Scheduled Tribe; Scheduled Caste; Other Backward Class. |
| **Household Characteristics** |  |  |
| Household size | num | Continuous household size. |
| Household headship (female) | binary | Categories: Yes (female head); No (non-female head). |
| Main source of drinking water | category | Categories: Pipe on premises; Other improved sources; Unimproved sources. |
| Time taken to fetch water | binary | Categories: Yes (less than 15 minutes); No (more than 15 minutes). |
| Home ownership | binary | Categories: Yes (own home); No (do not own home). |
| **Health Variables** |  |  |
| **Diagnosed chronic conditions or diseases** |  |  |
| Hypertension | binary | Categories: Yes (diagnosed); No (not diagnosed). |
| Diabetes | binary | Categories: Yes (diagnosed); No (not diagnosed). |
| Cancer | binary | Categories: Yes (diagnosed); No (not diagnosed). |
| Chronic lung disease | binary | Categories: Yes (diagnosed); No (not diagnosed). |
| Chronic heart diseases | binary | Categories: Yes (diagnosed); No (not diagnosed). |
| Stroke | binary | Categories: Yes (diagnosed); No (not diagnosed). |
| Bone/joint diseases | binary | Categories: Yes (diagnosed); No (not diagnosed). |
| Psychiatric problems | binary | Categories: Yes (diagnosed); No (not diagnosed). |
| High cholesterol | binary | Categories: Yes (diagnosed); No (not diagnosed). |
| **Urogenital** |  |  |
| Chronic Renal Failure | binary | Categories: Yes (diagnosed); No (not diagnosed). |
| Incontinence | binary | Categories: Yes (diagnosed); No (not diagnosed). |
| Kidney Stones | binary | Categories: Yes (diagnosed); No (not diagnosed). |
| BPH (Benign Prostatic Hyperplasia) | binary | Categories: Yes (diagnosed); No (not diagnosed). |
| **Immunization** |  |  |
| Influenza vaccine | binary | Categories: Yes (received); No (not received). |
| Pneumococcal vaccine | binary | Categories: Yes (received); No (not received). |
| Hepatitis B vaccine | binary | Categories: Yes (received); No (not received). |
| Typhoid vaccine | binary | Categories: Yes (received); No (not received). |
| Diphtheria and Tetanus (dT) | binary | Categories: Yes (received); No (not received). |
| **Symptom-based health** |  |  |
| Pain | binary | Categories: Yes (reported pain); No (no pain). |
| Sleep problem | binary | Categories: Yes (reported sleep problem); No (no sleep problem). |
| **Vector-borne diseases** |  |  |
| Malaria | binary | Categories: Yes (diagnosed); No (not diagnosed). |
| Dengue | binary | Categories: Yes (diagnosed); No (not diagnosed). |
| Chickungunya | binary | Categories: Yes (diagnosed); No (not diagnosed). |
| **Infectious diseases** |  |  |
| Tuberculosis | binary | Categories: Yes (diagnosed); No (not diagnosed). |
| Urinary tract infection | binary | Categories: Yes (diagnosed); No (not diagnosed). |
| **Others** |  |  |
| Overall Cognition score | binary | Categories: Lowest 10% (Yes); Not lowest 10% (No). |
| Depression (CESD measured) | binary | Categories: Yes (depressed); No (not depressed). |
| Self-rated general health | label | Categories: Very poor; Poor; Fair; Good; Very good. |
| **Functional Limitation Variables** |  |  |
| Vision | binary | Categories: Low vision (Yes); Not low vision (No). |
| Grip strength | num | Continuous measurement of grip strength. |
| Aids used | binary | Categories: Yes (using aids); No (not using aids). |
| ADL | binary | Categories: Yes (difficulty in at least one activity of daily living); No (no difficulty). |
| IADL | binary | Categories: Yes (difficulty in at least one instrumental activity of daily living); No (no difficulty). |
| Use of denture | binary | Categories: Yes (uses dentures); No (does not use dentures). |
| **Behavioral Risk Factors Variables** |  |  |
| Smoking | binary | Categories: Yes (currently smoking); No (not currently smoking). |
| Smokeless | binary | Categories: Yes (using smokeless tobacco); No (not using smokeless tobacco). |
| Alcohol | category | Categories: Lifetime abstainer; Infrequent non-heavy drinker; Frequent/heavy drinker. |
| Physically active | category | Categories: Low; Moderate; High. |
| Yoga/meditation/asana/pranayama | binary | Categories: Yes (engages more than once a week); No (does not engage regularly). |
| Food availability | binary | Categories: Yes (reported severe food constraints); No (no severe food constraints). |
| **Health Service Use and Health Insurance Variables** |  |  |
| Out-patient care | binary | Categories: Yes (any outpatient visit last year); No (none). |
| Inpatient care | binary | Categories: Yes (any inpatient visit last year); No (none). |
| Mean out-of-pocket expenditure on out-patient care in one year prior to the survey | num | Continuous variable representing expenditure. |
| Mean out-of-pocket expenditure on inpatient care in one year prior to the survey | num | Continuous variable representing expenditure. |
| Public Health insurance | binary | Categories: Yes (covered by public insurance); No (not covered). |
| Private Health insurance | binary | Categories: Yes (covered by private insurance); No (not covered). |
| **Family and Social Variables** |  |  |
| Making family decision | binary | Categories: Yes (involved in family decisions); No (not involved). |
| Look after grandchildren | binary | Categories: Yes (responsible for caregiving); No (not responsible). |
| Provide financial support | binary | Categories: Yes (provides financial help); No (does not provide help). |
| Received financial support | binary | Categories: Yes (receives financial help); No (does not receive help). |
| Having family members who are unable to carry out basic daily activities | binary | Categories: Yes (has dependent family members); No (none). |
| Membership in any organization | binary | Categories: Yes (member of organization); No (not a member). |
| Discrimination | binary | Categories: Yes (experienced discrimination); No (no experience of discrimination). |
| Life satisfaction | label | Categories: Low; Medium; High. |
| Receive social benefit | category | Number of benefits received. |
| **Outcome Variables** |  |  |
| BMI category | category | Categories: Underweight (BMI < 18.5); Normal (BMI 18.5–23); Overweight (BMI 23–25); Obesity (BMI > 25). |
| Waist circumference | binary | Categories: Male (> 94 cm); Female (> 80 cm). |

**Supplementary Table 3.** Comparison of Evaluation Metrics Across Different Machine Learning Models.

**(a) Underweight**
|  | **Accuracy**<br>**(95% CI)** | **Sensitivity**<br>**(95% CI)** | **Specificity**<br>**(95% CI)** | **Precision**<br>**(95% CI)** | **AUROC**<br>**(95% CI)** |
| --- | --- | --- | --- | --- | --- |
| **Logistic Regression** |  |  |  |  |  |
| Overall | 0.83 (0.82-0.83) | 0.23 (0.21-0.24) | 0.97 (0.96-0.97) | 0.62 (0.58-0.65) | 0.80 (0.79-0.81) |
| General | 0.88 (0.87-0.89) | 0.13 (0.10-0.17) | 0.99 (0.99-0.99) | 0.66 (0.55-0.76) | 0.80 (0.78-0.83) |
| Scheduled caste | 0.79 (0.78-0.81) | 0.34 (0.30-0.38) | 0.93 (0.92-0.94) | 0.60 (0.54-0.66) | 0.81 (0.78-0.83) |
| Scheduled tribe | 0.79 (0.77-0.81) | 0.25 (0.21-0.29) | 0.96 (0.95-0.97) | 0.67 (0.60-0.74) | 0.79 (0.77-0.81) |
| Other backward class | 0.81 (0.80-0.83) | 0.19 (0.17-0.22) | 0.97 (0.96-0.97) | 0.59 (0.53-0.65) | 0.79 (0.77-0.80) |
| MPCE Lowest | 0.73 (0.72-0.75) | 0.35 (0.31-0.39) | 0.91 (0.89-0.92) | 0.62 (0.57-0.66) | 0.75 (0.73-0.77) |
| MPCE Lower middle | 0.78 (0.77-0.80) | 0.23 (0.19-0.26) | 0.96 (0.95-0.97) | 0.62 (0.55-0.69) | 0.77 (0.75-0.79) |
| MPCE Middle | 0.81 (0.80-0.83) | 0.16 (0.12-0.19) | 0.97 (0.97-0.98) | 0.58 (0.49-0.67) | 0.79 (0.77-0.81) |
| MPCE Upper middle | 0.88 (0.87-0.89) | 0.13 (0.09-0.17) | 0.99 (0.98-0.99) | 0.64 (0.52-0.76) | 0.80 (0.77-0.82) |
| MPCE Highest | 0.92 (0.90-0.93) | 0.10 (0.06-0.15) | 1.00 (0.99-1.00) | 0.77 (0.60-0.92) | 0.80 (0.77-0.84) |
| **Random Forest** |  |  |  |  |  |
| Overall | 0.82 (0.81-0.82) | 0.09 (0.08-0.10) | 0.99 (0.98-0.99) | 0.61 (0.56-0.67) | 0.78 (0.77-0.79) |
| General | 0.88 (0.87-0.89) | 0.05 (0.03-0.07) | 1.00 (1.00-1.00) | 0.70 (0.53-0.86) | 0.78 (0.76-0.81) |
| Scheduled caste | 0.78 (0.77-0.80) | 0.16 (0.13-0.19) | 0.97 (0.96-0.98) | 0.64 (0.55-0.73) | 0.80 (0.78-0.83) |
| Scheduled tribe | 0.77 (0.75-0.79) | 0.09 (0.06-0.12) | 0.98 (0.98-0.99) | 0.64 (0.52-0.75) | 0.76 (0.73-0.78) |
| Other backward class | 0.80 (0.79-0.82) | 0.07 (0.05-0.09) | 0.99 (0.98-0.99) | 0.55 (0.46-0.64) | 0.76 (0.75-0.78) |
| MPCE Lowest | 0.72 (0.70-0.74) | 0.17 (0.14-0.19) | 0.96 (0.95-0.97) | 0.65 (0.57-0.72) | 0.74 (0.72-0.76) |
| MPCE Lower middle | 0.77 (0.75-0.78) | 0.08 (0.05-0.10) | 0.98 (0.97-0.99) | 0.55 (0.43-0.66) | 0.75 (0.73-0.77) |
| MPCE Middle | 0.81 (0.79-0.82) | 0.05 (0.03-0.07) | 0.99 (0.99-1.00) | 0.58 (0.42-0.74) | 0.77 (0.74-0.79) |
| MPCE Upper middle | 0.88 (0.86-0.89) | 0.05 (0.02-0.07) | 1.00 (0.99-1.00) | 0.63 (0.42-0.84) | 0.76 (0.74-0.79) |
| MPCE Highest | 0.91 (0.90-0.92) | 0.02 (0.00-0.04) | 1.00 (1.00-1.00) | 0.67 (0.20-1.00) | 0.78 (0.75-0.82) |
| **XGBoost** |  |  |  |  |  |
| Overall | 0.82 (0.81-0.83) | 0.26 (0.24-0.28) | 0.95 (0.95-0.96) | 0.57 (0.54-0.59) | 0.79 (0.78-0.80) |
| General | 0.88 (0.87-0.89) | 0.15 (0.12-0.19) | 0.98 (0.97-0.98) | 0.51 (0.42-0.59) | 0.79 (0.77-0.82) |
| Scheduled caste | 0.80 (0.78-0.82) | 0.39 (0.35-0.44) | 0.92 (0.91-0.94) | 0.61 (0.56-0.67) | 0.80 (0.78-0.82) |
| Scheduled tribe | 0.78 (0.77-0.80) | 0.27 (0.23-0.32) | 0.95 (0.93-0.96) | 0.61 (0.54-0.68) | 0.77 (0.74-0.79) |
| Other backward class | 0.81 (0.79-0.82) | 0.23 (0.20-0.26) | 0.95 (0.94-0.96) | 0.52 (0.47-0.57) | 0.78 (0.76-0.79) |
| MPCE Lowest | 0.73 (0.71-0.75) | 0.37 (0.34-0.41) | 0.88 (0.87-0.90) | 0.58 (0.54-0.63) | 0.73 (0.71-0.75) |
| MPCE Lower middle | 0.78 (0.77-0.80) | 0.27 (0.23-0.31) | 0.94 (0.93-0.95) | 0.59 (0.53-0.65) | 0.76 (0.74-0.78) |
| MPCE Middle | 0.81 (0.80-0.83) | 0.20 (0.17-0.24) | 0.96 (0.95-0.97) | 0.55 (0.47-0.62) | 0.79 (0.76-0.81) |
| MPCE Upper middle | 0.87 (0.86-0.88) | 0.15 (0.11-0.20) | 0.97 (0.97-0.98) | 0.45 (0.35-0.55) | 0.77 (0.74-0.80) |
| MPCE Highest | 0.91 (0.90-0.92) | 0.14 (0.09-0.19) | 0.99 (0.99-0.99) | 0.58 (0.43-0.72) | 0.79 (0.76-0.83) |

| **Gradient Boosting** |  |  |  |  |  |
| --- | --- | --- | --- | --- | --- |
| Overall | 0.82 (0.82-0.83) | 0.16 (0.15-0.18) | 0.98 (0.98-0.98) | 0.65 (0.61-0.69) | 0.80 (0.79-0.81) |
| General | 0.88 (0.87-0.89) | 0.11 (0.08-0.14) | 0.99 (0.99-1.00) | 0.71 (0.59-0.82) | 0.80 (0.78-0.82) |
| Scheduled caste | 0.79 (0.77-0.81) | 0.26 (0.22-0.30) | 0.95 (0.94-0.96) | 0.63 (0.56-0.70) | 0.81 (0.79-0.83) |
| Scheduled tribe | 0.78 (0.76-0.80) | 0.17 (0.14-0.21) | 0.98 (0.97-0.98) | 0.69 (0.60-0.77) | 0.79 (0.76-0.81) |
| Other backward class | 0.81 (0.80-0.82) | 0.13 (0.11-0.16) | 0.98 (0.98-0.98) | 0.63 (0.55-0.70) | 0.79 (0.77-0.80) |
| MPCE Lowest | 0.74 (0.72-0.75) | 0.29 (0.25-0.32) | 0.93 (0.92-0.95) | 0.65 (0.60-0.71) | 0.75 (0.73-0.77) |
| MPCE Lower middle | 0.78 (0.76-0.79) | 0.13 (0.11-0.16) | 0.97 (0.97-0.98) | 0.62 (0.53-0.71) | 0.77 (0.75-0.79) |
| MPCE Middle | 0.82 (0.80-0.83) | 0.11 (0.08-0.14) | 0.99 (0.98-0.99) | 0.68 (0.56-0.78) | 0.79 (0.77-0.81) |
| MPCE Upper middle | 0.88 (0.87-0.89) | 0.09 (0.06-0.12) | 0.99 (0.99-0.99) | 0.58 (0.43-0.73) | 0.78 (0.76-0.81) |
| MPCE Highest | 0.91 (0.90-0.93) | 0.08 (0.04-0.12) | 1.00 (1.00-1.00) | 0.79 (0.58-0.95) | 0.81 (0.77-0.84) |
| **LightGBM** |  |  |  |  |  |
| Overall | 0.82 (0.82-0.83) | 0.22 (0.20-0.23) | 0.97 (0.96-0.97) | 0.60 (0.57-0.63) | 0.80 (0.79-0.81) |
| General | 0.88 (0.87-0.89) | 0.14 (0.10-0.17) | 0.99 (0.98-0.99) | 0.60 (0.49-0.71) | 0.80 (0.78-0.82) |
| Scheduled caste | 0.79 (0.78-0.81) | 0.31 (0.27-0.36) | 0.94 (0.93-0.95) | 0.61 (0.55-0.67) | 0.81 (0.79-0.83) |
| Scheduled tribe | 0.79 (0.77-0.81) | 0.26 (0.22-0.30) | 0.95 (0.94-0.96) | 0.63 (0.56-0.69) | 0.78 (0.76-0.81) |
| Other backward class | 0.81 (0.80-0.82) | 0.18 (0.15-0.21) | 0.97 (0.96-0.97) | 0.57 (0.51-0.63) | 0.79 (0.77-0.80) |
| MPCE Lowest | 0.73 (0.71-0.75) | 0.32 (0.28-0.36) | 0.91 (0.89-0.92) | 0.61 (0.56-0.66) | 0.75 (0.73-0.77) |
| MPCE Lower middle | 0.78 (0.77-0.80) | 0.23 (0.20-0.27) | 0.95 (0.94-0.96) | 0.61 (0.54-0.67) | 0.77 (0.75-0.79) |
| MPCE Middle | 0.81 (0.79-0.83) | 0.16 (0.13-0.20) | 0.97 (0.96-0.98) | 0.55 (0.47-0.64) | 0.79 (0.77-0.81) |
| MPCE Upper middle | 0.88 (0.87-0.89) | 0.14 (0.10-0.18) | 0.98 (0.98-0.99) | 0.57 (0.46-0.68) | 0.79 (0.76-0.82) |
| MPCE Highest | 0.91 (0.90-0.93) | 0.08 (0.04-0.11) | 1.00 (0.99-1.00) | 0.71 (0.50-0.90) | 0.81 (0.78-0.84) |
| **Deep Neural Networks (DNN)** |  |  |  |  |  |
| Overall | 0.79 (0.79-0.80) | 0.37 (0.35-0.39) | 0.89 (0.89-0.90) | 0.45 (0.43-0.47) | 0.76 (0.75-0.77) |
| General | 0.86 (0.85-0.87) | 0.26 (0.21-0.30) | 0.95 (0.94-0.95) | 0.40 (0.34-0.47) | 0.75 (0.73-0.78) |
| Scheduled caste | 0.76 (0.75-0.78) | 0.48 (0.43-0.53) | 0.85 (0.83-0.87) | 0.50 (0.45-0.54) | 0.78 (0.75-0.80) |
| Scheduled tribe | 0.75 (0.73-0.77) | 0.43 (0.38-0.47) | 0.85 (0.83-0.87) | 0.47 (0.42-0.52) | 0.74 (0.72-0.77) |
| Other backward class | 0.78 (0.76-0.79) | 0.34 (0.30-0.37) | 0.89 (0.87-0.90) | 0.42 (0.38-0.46) | 0.74 (0.72-0.75) |
| MPCE Lowest | 0.68 (0.66-0.70) | 0.48 (0.44-0.52) | 0.77 (0.75-0.79) | 0.48 (0.44-0.52) | 0.70 (0.68-0.72) |
| MPCE Lower middle | 0.76 (0.74-0.78) | 0.40 (0.36-0.45) | 0.87 (0.85-0.88) | 0.48 (0.44-0.53) | 0.74 (0.71-0.76) |
| MPCE Middle | 0.79 (0.77-0.80) | 0.31 (0.27-0.35) | 0.90 (0.89-0.91) | 0.43 (0.38-0.48) | 0.73 (0.71-0.76) |
| MPCE Upper middle | 0.84 (0.83-0.86) | 0.25 (0.20-0.30) | 0.93 (0.92-0.94) | 0.33 (0.27-0.40) | 0.74 (0.72-0.77) |
| MPCE Highest | 0.90 (0.89-0.92) | 0.24 (0.18-0.30) | 0.97 (0.96-0.98) | 0.44 (0.34-0.53) | 0.76 (0.72-0.79) |
| **Fully Convolutional Networks (FCN)** |  |  |  |  |  |
| Overall | 0.77 (0.76-0.78) | 0.32 (0.30-0.34) | 0.88 (0.87-0.89) | 0.38 (0.36-0.41) | 0.71 (0.70-0.72) |
| General | 0.84 (0.82-0.85) | 0.21 (0.17-0.25) | 0.93 (0.92-0.94) | 0.29 (0.23-0.34) | 0.70 (0.67-0.72) |
| Scheduled caste | 0.74 (0.72-0.75) | 0.39 (0.34-0.43) | 0.84 (0.82-0.86) | 0.43 (0.38-0.47) | 0.72 (0.69-0.74) |
| Scheduled tribe | 0.73 (0.71-0.75) | 0.36 (0.32-0.41) | 0.84 (0.82-0.86) | 0.42 (0.37-0.47) | 0.69 (0.66-0.71) |
| Other backward class | 0.76 (0.75-0.78) | 0.30 (0.27-0.33) | 0.88 (0.87-0.89) | 0.38 (0.35-0.42) | 0.70 (0.68-0.72) |
| MPCE Lowest | 0.66 (0.64-0.68) | 0.39 (0.36-0.43) | 0.78 (0.76-0.80) | 0.44 (0.40-0.48) | 0.66 (0.63-0.68) |
| MPCE Lower middle | 0.74 (0.72-0.75) | 0.33 (0.29-0.37) | 0.86 (0.85-0.88) | 0.42 (0.38-0.47) | 0.68 (0.65-0.70) |
| MPCE Middle | 0.77 (0.75-0.78) | 0.31 (0.27-0.35) | 0.88 (0.86-0.89) | 0.38 (0.33-0.43) | 0.69 (0.66-0.72) |
| MPCE Upper middle | 0.82 (0.80-0.83) | 0.23 (0.18-0.28) | 0.90 (0.89-0.92) | 0.25 (0.20-0.31) | 0.68 (0.65-0.71) |
| MPCE Highest | 0.88 (0.87-0.89) | 0.16 (0.12-0.22) | 0.95 (0.94-0.96) | 0.25 (0.18-0.33) | 0.71 (0.67-0.74) |

**(b) Overweight/Obesity**
|  | Accuracy<br>(95% CI) | Sensitivity<br>(95% CI) | Specificity<br>(95% CI) | Precision<br>(95% CI) | AUROC<br>(95% CI) |
| --- | --- | --- | --- | --- | --- |
| **Logistic Regression** |  |  |  |  |  |
| Overall | 0.73 (0.72-0.74) | 0.65 (0.64-0.67) | 0.79 (0.78-0.80) | 0.71 (0.69-0.72) | 0.80 (0.79-0.81) |
| General | 0.71 (0.69-0.73) | 0.78 (0.76-0.80) | 0.62 (0.59-0.64) | 0.73 (0.70-0.75) | 0.78 (0.76-0.79) |
| Scheduled caste | 0.75 (0.73-0.77) | 0.45 (0.40-0.48) | 0.89 (0.87-0.90) | 0.64 (0.59-0.68) | 0.78 (0.76-0.80) |
| Scheduled tribe | 0.73 (0.71-0.75) | 0.53 (0.50-0.57) | 0.85 (0.83-0.87) | 0.69 (0.65-0.73) | 0.79 (0.77-0.81) |
| Other backward class | 0.73 (0.72-0.74) | 0.65 (0.63-0.67) | 0.79 (0.77-0.81) | 0.71 (0.69-0.73) | 0.80 (0.78-0.81) |
| MPCE Lowest | 0.78 (0.76-0.79) | 0.34 (0.30-0.38) | 0.93 (0.92-0.95) | 0.65 (0.60-0.70) | 0.78 (0.76-0.81) |
| MPCE Lower middle | 0.72 (0.71-0.74) | 0.49 (0.45-0.52) | 0.86 (0.84-0.88) | 0.67 (0.63-0.71) | 0.77 (0.75-0.79) |
| MPCE Middle | 0.72 (0.71-0.74) | 0.64 (0.61-0.67) | 0.79 (0.76-0.81) | 0.69 (0.66-0.72) | 0.79 (0.77-0.81) |
| MPCE Upper middle | 0.69 (0.67-0.71) | 0.71 (0.69-0.74) | 0.66 (0.63-0.69) | 0.71 (0.68-0.74) | 0.76 (0.74-0.78) |
| MPCE Highest | 0.72 (0.70-0.74) | 0.85 (0.83-0.87) | 0.51 (0.48-0.55) | 0.74 (0.72-0.76) | 0.77 (0.75-0.79) |
| **Random Forest** |  |  |  |  |  |
| Overall | 0.72 (0.71-0.73) | 0.63 (0.62-0.64) | 0.79 (0.78-0.80) | 0.70 (0.69-0.71) | 0.79 (0.78-0.79) |
| General | 0.70 (0.68-0.71) | 0.74 (0.72-0.76) | 0.65 (0.62-0.67) | 0.73 (0.71-0.75) | 0.76 (0.74-0.78) |
| Scheduled caste | 0.74 (0.72-0.76) | 0.44 (0.40-0.47) | 0.87 (0.85-0.89) | 0.61 (0.56-0.65) | 0.76 (0.74-0.79) |
| Scheduled tribe | 0.73 (0.71-0.75) | 0.55 (0.52-0.59) | 0.84 (0.82-0.86) | 0.68 (0.65-0.72) | 0.78 (0.76-0.80) |
| Other backward class | 0.72 (0.71-0.73) | 0.63 (0.61-0.65) | 0.79 (0.78-0.81) | 0.70 (0.68-0.73) | 0.78 (0.77-0.80) |
| MPCE Lowest | 0.77 (0.75-0.79) | 0.33 (0.29-0.37) | 0.93 (0.92-0.95) | 0.64 (0.59-0.69) | 0.77 (0.74-0.79) |
| MPCE Lower middle | 0.72 (0.70-0.74) | 0.48 (0.45-0.52) | 0.86 (0.84-0.87) | 0.66 (0.62-0.69) | 0.75 (0.73-0.77) |
| MPCE Middle | 0.71 (0.69-0.73) | 0.61 (0.58-0.64) | 0.79 (0.76-0.81) | 0.68 (0.65-0.71) | 0.78 (0.76-0.79) |
| MPCE Upper middle | 0.69 (0.67-0.71) | 0.69 (0.67-0.72) | 0.69 (0.66-0.71) | 0.72 (0.70-0.75) | 0.75 (0.73-0.77) |
| MPCE Highest | 0.70 (0.68-0.72) | 0.81 (0.79-0.83) | 0.52 (0.49-0.55) | 0.73 (0.71-0.76) | 0.74 (0.72-0.76) |
| **XGBoost** |  |  |  |  |  |
| Overall | 0.72 (0.71-0.73) | 0.65 (0.64-0.67) | 0.78 (0.76-0.79) | 0.70 (0.68-0.71) | 0.79 (0.78-0.80) |
| General | 0.70 (0.68-0.71) | 0.77 (0.75-0.79) | 0.61 (0.58-0.64) | 0.72 (0.70-0.74) | 0.77 (0.75-0.78) |
| Scheduled caste | 0.74 (0.72-0.76) | 0.48 (0.44-0.52) | 0.86 (0.84-0.88) | 0.61 (0.56-0.65) | 0.77 (0.75-0.79) |
| Scheduled tribe | 0.72 (0.70-0.74) | 0.53 (0.50-0.57) | 0.83 (0.81-0.85) | 0.66 (0.62-0.70) | 0.78 (0.76-0.80) |
| Other backward class | 0.73 (0.72-0.74) | 0.65 (0.63-0.67) | 0.79 (0.77-0.81) | 0.71 (0.69-0.73) | 0.79 (0.78-0.80) |
| MPCE Lowest | 0.77 (0.75-0.79) | 0.38 (0.34-0.42) | 0.91 (0.90-0.92) | 0.61 (0.56-0.65) | 0.78 (0.76-0.80) |
| MPCE Lower middle | 0.73 (0.71-0.74) | 0.52 (0.48-0.55) | 0.85 (0.83-0.86) | 0.66 (0.62-0.70) | 0.76 (0.74-0.78) |
| MPCE Middle | 0.72 (0.70-0.74) | 0.63 (0.60-0.66) | 0.79 (0.76-0.81) | 0.68 (0.65-0.71) | 0.78 (0.76-0.80) |
| MPCE Upper middle | 0.69 (0.67-0.71) | 0.71 (0.69-0.74) | 0.66 (0.63-0.69) | 0.71 (0.68-0.73) | 0.76 (0.74-0.78) |
| MPCE Highest | 0.70 (0.68-0.72) | 0.82 (0.80-0.84) | 0.52 (0.48-0.55) | 0.73 (0.71-0.76) | 0.75 (0.73-0.77) |
| **Gradient Boosting** |  |  |  |  |  |
| Overall | 0.72 (0.71-0.73) | 0.64 (0.62-0.65) | 0.79 (0.78-0.80) | 0.71 (0.69-0.72) | 0.79 (0.79-0.80) |
| General | 0.70 (0.69-0.72) | 0.74 (0.73-0.76) | 0.65 (0.62-0.68) | 0.74 (0.71-0.76) | 0.77 (0.75-0.79) |
| Scheduled caste | 0.74 (0.73-0.76) | 0.45 (0.41-0.48) | 0.88 (0.86-0.90) | 0.63 (0.58-0.67) | 0.78 (0.76-0.80) |
| Scheduled tribe | 0.73 (0.71-0.74) | 0.53 (0.50-0.57) | 0.84 (0.82-0.86) | 0.68 (0.64-0.71) | 0.79 (0.77-0.81) |
| Other backward class | 0.72 (0.71-0.74) | 0.64 (0.62-0.66) | 0.79 (0.78-0.81) | 0.71 (0.68-0.73) | 0.79 (0.78-0.80) |
| MPCE Lowest | 0.77 (0.75-0.79) | 0.34 (0.30-0.38) | 0.93 (0.91-0.94) | 0.63 (0.58-0.68) | 0.78 (0.76-0.80) |
| MPCE Lower middle | 0.72 (0.70-0.73) | 0.47 (0.44-0.51) | 0.85 (0.84-0.87) | 0.65 (0.61-0.69) | 0.77 (0.75-0.79) |
| MPCE Middle | 0.72 (0.70-0.74) | 0.62 (0.59-0.65) | 0.79 (0.77-0.81) | 0.69 (0.66-0.72) | 0.78 (0.76-0.80) |
| MPCE Upper middle | 0.69 (0.67-0.71) | 0.69 (0.66-0.72) | 0.69 (0.66-0.72) | 0.72 (0.70-0.74) | 0.76 (0.74-0.78) |
| MPCE Highest | 0.72 (0.70-0.74) | 0.83 (0.81-0.85) | 0.54 (0.51-0.57) | 0.75 (0.72-0.77) | 0.76 (0.74-0.78) |
| **LightGBM** |  |  |  |  |  |
| Overall | 0.72 (0.71-0.73) | 0.65 (0.64-0.67) | 0.78 (0.77-0.79) | 0.70 (0.68-0.71) | 0.80 (0.79-0.81) |
| General | 0.70 (0.68-0.71) | 0.76 (0.74-0.78) | 0.61 (0.59-0.64) | 0.72 (0.70-0.74) | 0.77 (0.76-0.79) |
| Scheduled caste | 0.74 (0.72-0.76) | 0.47 (0.43-0.51) | 0.87 (0.85-0.89) | 0.62 (0.57-0.66) | 0.79 (0.77-0.81) |
| Scheduled tribe | 0.73 (0.71-0.75) | 0.55 (0.51-0.58) | 0.84 (0.82-0.86) | 0.68 (0.64-0.71) | 0.79 (0.77-0.81) |
| Other backward class | 0.73 (0.72-0.74) | 0.66 (0.64-0.68) | 0.78 (0.77-0.80) | 0.70 (0.68-0.73) | 0.80 (0.78-0.81) |
| MPCE Lowest | 0.78 (0.76-0.79) | 0.37 (0.33-0.41) | 0.92 (0.91-0.94) | 0.63 (0.58-0.69) | 0.78 (0.76-0.81) |
| MPCE Lower middle | 0.72 (0.70-0.74) | 0.52 (0.48-0.55) | 0.84 (0.82-0.86) | 0.65 (0.61-0.68) | 0.76 (0.75-0.79) |
| MPCE Middle | 0.71 (0.69-0.73) | 0.63 (0.60-0.66) | 0.78 (0.75-0.80) | 0.67 (0.65-0.70) | 0.78 (0.76-0.80) |
| MPCE Upper middle | 0.69 (0.67-0.71) | 0.71 (0.68-0.73) | 0.67 (0.65-0.70) | 0.72 (0.69-0.74) | 0.76 (0.74-0.78) |
| MPCE Highest | 0.71 (0.69-0.73) | 0.84 (0.82-0.86) | 0.51 (0.47-0.54) | 0.73 (0.71-0.76) | 0.77 (0.75-0.79) |
| **Deep Neural Networks (DNN)** |  |  |  |  |  |
| Overall | 0.70 (0.69-0.71) | 0.68 (0.66-0.69) | 0.71 (0.70-0.72) | 0.65 (0.64-0.66) | 0.76 (0.76-0.77) |
| General | 0.68 (0.67-0.70) | 0.78 (0.76-0.80) | 0.56 (0.54-0.59) | 0.70 (0.68-0.72) | 0.74 (0.72-0.76) |
| Scheduled caste | 0.72 (0.70-0.74) | 0.51 (0.47-0.55) | 0.81 (0.79-0.83) | 0.55 (0.51-0.59) | 0.75 (0.73-0.77) |
| Scheduled tribe | 0.70 (0.68-0.72) | 0.58 (0.54-0.61) | 0.77 (0.74-0.79) | 0.61 (0.57-0.64) | 0.75 (0.73-0.77) |
| Other backward class | 0.70 (0.68-0.71) | 0.67 (0.65-0.69) | 0.71 (0.70-0.73) | 0.65 (0.62-0.67) | 0.76 (0.75-0.78) |
| MPCE Lowest | 0.75 (0.73-0.77) | 0.44 (0.40-0.48) | 0.86 (0.85-0.88) | 0.54 (0.50-0.58) | 0.75 (0.72-0.77) |
| MPCE Lower middle | 0.69 (0.67-0.71) | 0.56 (0.53-0.59) | 0.76 (0.74-0.79) | 0.58 (0.54-0.61) | 0.73 (0.71-0.75) |
| MPCE Middle | 0.68 (0.66-0.70) | 0.65 (0.63-0.68) | 0.70 (0.68-0.73) | 0.62 (0.59-0.65) | 0.75 (0.73-0.77) |
| MPCE Upper middle | 0.67 (0.65-0.69) | 0.73 (0.71-0.76) | 0.60 (0.56-0.63) | 0.68 (0.65-0.70) | 0.74 (0.72-0.76) |
| MPCE Highest | 0.69 (0.67-0.71) | 0.82 (0.80-0.84) | 0.49 (0.46-0.53) | 0.72 (0.70-0.75) | 0.73 (0.70-0.75) |
| **Fully Convolutional Networks (FCN)** |  |  |  |  |  |
| Overall | 0.66 (0.65-0.67) | 0.63 (0.61-0.64) | 0.68 (0.67-0.69) | 0.61 (0.59-0.62) | 0.71 (0.70-0.72) |
| General | 0.66 (0.64-0.67) | 0.73 (0.71-0.75) | 0.57 (0.54-0.59) | 0.69 (0.66-0.71) | 0.70 (0.68-0.72) |
| Scheduled caste | 0.67 (0.65-0.69) | 0.47 (0.43-0.50) | 0.76 (0.74-0.79) | 0.47 (0.43-0.51) | 0.68 (0.66-0.71) |
| Scheduled tribe | 0.66 (0.64-0.68) | 0.54 (0.50-0.58) | 0.73 (0.71-0.76) | 0.55 (0.52-0.59) | 0.68 (0.66-0.71) |
| Other backward class | 0.65 (0.64-0.67) | 0.62 (0.60-0.64) | 0.67 (0.65-0.69) | 0.60 (0.58-0.62) | 0.70 (0.68-0.72) |
| MPCE Lowest | 0.71 (0.69-0.72) | 0.44 (0.41-0.48) | 0.80 (0.78-0.82) | 0.45 (0.40-0.48) | 0.69 (0.66-0.72) |
| MPCE Lower middle | 0.66 (0.64-0.68) | 0.54 (0.51-0.58) | 0.74 (0.71-0.76) | 0.54 (0.51-0.57) | 0.68 (0.66-0.71) |
| MPCE Middle | 0.64 (0.62-0.66) | 0.59 (0.56-0.62) | 0.67 (0.65-0.70) | 0.57 (0.54-0.60) | 0.68 (0.66-0.70) |
| MPCE Upper middle | 0.62 (0.61-0.64) | 0.65 (0.63-0.68) | 0.59 (0.56-0.62) | 0.65 (0.62-0.68) | 0.67 (0.65-0.69) |
| MPCE Highest | 0.66 (0.64-0.68) | 0.77 (0.74-0.79) | 0.48 (0.45-0.52) | 0.71 (0.68-0.73) | 0.69 (0.66-0.71) |

**(c) High Waist Circumference**
|  | **Accuracy**<br>**(95% CI)** | **Sensitivity**<br>**(95% CI)** | **Specificity**<br>**(95% CI)** | **Precision**<br>**(95% CI)** | **AUROC**<br>**(95% CI)** |
| --- | --- | --- | --- | --- | --- |
| **Logistic Regression** |  |  |  |  |  |
| Overall | 0.75 (0.74-0.76) | 0.71 (0.70-0.72) | 0.79 (0.78-0.80) | 0.74 (0.73-0.75) | 0.84 (0.83-0.84) |
| General | 0.75 (0.74-0.77) | 0.81 (0.79-0.83) | 0.66 (0.63-0.69) | 0.77 (0.76-0.79) | 0.83 (0.82-0.85) |
| Scheduled caste | 0.77 (0.75-0.79) | 0.51 (0.47-0.55) | 0.89 (0.87-0.90) | 0.67 (0.63-0.71) | 0.82 (0.80-0.84) |
| Scheduled tribe | 0.74 (0.72-0.76) | 0.64 (0.61-0.67) | 0.83 (0.80-0.85) | 0.74 (0.70-0.77) | 0.83 (0.81-0.85) |
| Other backward class | 0.75 (0.73-0.76) | 0.71 (0.69-0.73) | 0.78 (0.76-0.79) | 0.72 (0.70-0.74) | 0.83 (0.82-0.84) |
| MPCE Lowest | 0.77 (0.75-0.79) | 0.51 (0.47-0.55) | 0.89 (0.87-0.90) | 0.67 (0.63-0.72) | 0.82 (0.80-0.83) |
| MPCE Lower middle | 0.75 (0.73-0.76) | 0.62 (0.59-0.65) | 0.83 (0.81-0.85) | 0.69 (0.66-0.72) | 0.82 (0.80-0.83) |
| MPCE Middle | 0.75 (0.73-0.76) | 0.72 (0.69-0.75) | 0.77 (0.74-0.79) | 0.72 (0.69-0.74) | 0.84 (0.82-0.85) |
| MPCE Upper middle | 0.75 (0.73-0.77) | 0.75 (0.72-0.77) | 0.75 (0.72-0.77) | 0.78 (0.76-0.81) | 0.83 (0.81-0.84) |
| MPCE Highest | 0.74 (0.72-0.76) | 0.83 (0.81-0.85) | 0.61 (0.58-0.64) | 0.77 (0.74-0.79) | 0.83 (0.81-0.85) |
| **Random Forest** |  |  |  |  |  |
| Overall | 0.75 (0.74-0.76) | 0.69 (0.68-0.70) | 0.80 (0.79-0.81) | 0.75 (0.73-0.76) | 0.83 (0.82-0.84) |
| General | 0.75 (0.73-0.77) | 0.78 (0.76-0.80) | 0.71 (0.69-0.74) | 0.79 (0.78-0.81) | 0.83 (0.81-0.84) |
| Scheduled caste | 0.77 (0.75-0.79) | 0.52 (0.48-0.56) | 0.89 (0.87-0.90) | 0.67 (0.63-0.71) | 0.82 (0.80-0.84) |
| Scheduled tribe | 0.74 (0.73-0.76) | 0.64 (0.60-0.67) | 0.83 (0.81-0.85) | 0.74 (0.71-0.77) | 0.82 (0.80-0.84) |
| Other backward class | 0.74 (0.73-0.75) | 0.68 (0.66-0.70) | 0.79 (0.77-0.80) | 0.72 (0.70-0.74) | 0.82 (0.80-0.83) |
| MPCE Lowest | 0.77 (0.75-0.79) | 0.50 (0.46-0.53) | 0.89 (0.88-0.91) | 0.67 (0.63-0.71) | 0.81 (0.79-0.83) |
| MPCE Lower middle | 0.74 (0.73-0.76) | 0.61 (0.58-0.65) | 0.83 (0.81-0.85) | 0.69 (0.66-0.72) | 0.81 (0.79-0.82) |
| MPCE Middle | 0.74 (0.72-0.76) | 0.68 (0.65-0.71) | 0.79 (0.77-0.81) | 0.73 (0.70-0.75) | 0.83 (0.81-0.84) |
| MPCE Upper middle | 0.74 (0.72-0.76) | 0.72 (0.69-0.74) | 0.76 (0.74-0.79) | 0.79 (0.76-0.81) | 0.82 (0.80-0.84) |
| MPCE Highest | 0.75 (0.73-0.77) | 0.81 (0.79-0.83) | 0.66 (0.63-0.69) | 0.78 (0.76-0.81) | 0.82 (0.80-0.84) |
| **XGBoost** |  |  |  |  |  |
| Overall | 0.75 (0.74-0.76) | 0.71 (0.69-0.72) | 0.79 (0.78-0.80) | 0.74 (0.73-0.75) | 0.83 (0.82-0.84) |
| General | 0.75 (0.74-0.77) | 0.80 (0.78-0.82) | 0.68 (0.65-0.71) | 0.78 (0.76-0.80) | 0.83 (0.82-0.84) |
| Scheduled caste | 0.76 (0.75-0.78) | 0.52 (0.48-0.56) | 0.88 (0.86-0.89) | 0.65 (0.61-0.69) | 0.81 (0.79-0.83) |
| Scheduled tribe | 0.74 (0.72-0.76) | 0.64 (0.61-0.68) | 0.82 (0.80-0.84) | 0.73 (0.69-0.76) | 0.82 (0.81-0.84) |
| Other backward class | 0.74 (0.73-0.76) | 0.70 (0.68-0.72) | 0.78 (0.76-0.79) | 0.72 (0.70-0.74) | 0.82 (0.81-0.83) |
| MPCE Lowest | 0.78 (0.76-0.80) | 0.54 (0.51-0.58) | 0.88 (0.87-0.90) | 0.68 (0.64-0.72) | 0.82 (0.80-0.84) |
| MPCE Lower middle | 0.74 (0.73-0.76) | 0.62 (0.59-0.66) | 0.82 (0.80-0.84) | 0.69 (0.65-0.72) | 0.81 (0.79-0.83) |
| MPCE Middle | 0.73 (0.72-0.75) | 0.69 (0.66-0.72) | 0.77 (0.74-0.79) | 0.71 (0.68-0.74) | 0.82 (0.81-0.84) |
| MPCE Upper middle | 0.74 (0.72-0.75) | 0.74 (0.71-0.76) | 0.74 (0.71-0.76) | 0.77 (0.75-0.80) | 0.82 (0.81-0.84) |
| MPCE Highest | 0.75 (0.73-0.77) | 0.82 (0.80-0.84) | 0.64 (0.61-0.67) | 0.78 (0.76-0.80) | 0.82 (0.80-0.84) |
| **Gradient Boosting** |  |  |  |  |  |
| Overall | 0.75 (0.75-0.76) | 0.70 (0.69-0.72) | 0.80 (0.79-0.81) | 0.75 (0.73-0.76) | 0.84 (0.83-0.84) |
| General | 0.75 (0.74-0.77) | 0.78 (0.76-0.81) | 0.71 (0.68-0.73) | 0.79 (0.77-0.81) | 0.83 (0.82-0.85) |
| Scheduled caste | 0.77 (0.76-0.79) | 0.53 (0.49-0.57) | 0.88 (0.87-0.90) | 0.67 (0.63-0.71) | 0.82 (0.80-0.84) |
| Scheduled tribe | 0.75 (0.73-0.77) | 0.66 (0.63-0.70) | 0.82 (0.79-0.84) | 0.73 (0.70-0.76) | 0.83 (0.81-0.85) |
| Other backward class | 0.74 (0.73-0.76) | 0.70 (0.68-0.72) | 0.78 (0.76-0.80) | 0.72 (0.70-0.74) | 0.82 (0.81-0.84) |
| MPCE Lowest | 0.77 (0.76-0.79) | 0.49 (0.45-0.53) | 0.90 (0.88-0.92) | 0.69 (0.65-0.73) | 0.81 (0.80-0.83) |
| MPCE Lower middle | 0.75 (0.73-0.77) | 0.64 (0.61-0.68) | 0.81 (0.79-0.84) | 0.69 (0.65-0.71) | 0.82 (0.80-0.83) |
| MPCE Middle | 0.75 (0.73-0.77) | 0.71 (0.68-0.73) | 0.78 (0.76-0.81) | 0.73 (0.70-0.76) | 0.83 (0.82-0.85) |
| MPCE Upper middle | 0.75 (0.73-0.76) | 0.73 (0.71-0.76) | 0.76 (0.74-0.79) | 0.79 (0.77-0.82) | 0.83 (0.81-0.84) |
| MPCE Highest | 0.75 (0.73-0.77) | 0.82 (0.80-0.84) | 0.64 (0.60-0.67) | 0.78 (0.75-0.80) | 0.83 (0.81-0.84) |
| **LightGBM** |  |  |  |  |  |
| Overall | 0.75 (0.75-0.76) | 0.71 (0.70-0.72) | 0.79 (0.78-0.80) | 0.75 (0.73-0.76) | 0.84 (0.83-0.85) |
| General | 0.75 (0.74-0.77) | 0.80 (0.78-0.81) | 0.69 (0.67-0.72) | 0.79 (0.77-0.80) | 0.83 (0.82-0.85) |
| Scheduled caste | 0.77 (0.75-0.79) | 0.53 (0.49-0.56) | 0.88 (0.86-0.90) | 0.66 (0.62-0.70) | 0.83 (0.81-0.85) |
| Scheduled tribe | 0.76 (0.74-0.78) | 0.66 (0.63-0.69) | 0.84 (0.82-0.86) | 0.76 (0.72-0.79) | 0.83 (0.81-0.85) |
| Other backward class | 0.75 (0.73-0.76) | 0.71 (0.68-0.73) | 0.78 (0.76-0.80) | 0.72 (0.70-0.74) | 0.83 (0.82-0.84) |
| MPCE Lowest | 0.78 (0.76-0.79) | 0.52 (0.48-0.56) | 0.89 (0.88-0.91) | 0.68 (0.64-0.73) | 0.82 (0.80-0.84) |
| MPCE Lower middle | 0.75 (0.74-0.77) | 0.64 (0.61-0.67) | 0.82 (0.80-0.84) | 0.70 (0.67-0.73) | 0.82 (0.80-0.84) |
| MPCE Middle | 0.75 (0.73-0.76) | 0.70 (0.68-0.73) | 0.78 (0.76-0.80) | 0.72 (0.70-0.75) | 0.83 (0.82-0.85) |
| MPCE Upper middle | 0.75 (0.73-0.76) | 0.74 (0.71-0.76) | 0.76 (0.73-0.78) | 0.79 (0.76-0.81) | 0.83 (0.81-0.85) |
| MPCE Highest | 0.75 (0.73-0.77) | 0.83 (0.80-0.85) | 0.63 (0.60-0.66) | 0.78 (0.75-0.80) | 0.83 (0.81-0.85) |
| **Deep Neural Networks (DNN)** |  |  |  |  |  |
| Overall | 0.73 (0.73-0.74) | 0.68 (0.67-0.69) | 0.78 (0.77-0.79) | 0.73 (0.71-0.74) | 0.81 (0.80-0.82) |
| General | 0.73 (0.72-0.75) | 0.77 (0.75-0.79) | 0.67 (0.65-0.70) | 0.77 (0.75-0.79) | 0.80 (0.79-0.82) |
| Scheduled caste | 0.76 (0.74-0.78) | 0.52 (0.48-0.56) | 0.87 (0.85-0.89) | 0.64 (0.60-0.69) | 0.80 (0.78-0.82) |
| Scheduled tribe | 0.73 (0.71-0.75) | 0.61 (0.58-0.65) | 0.81 (0.79-0.83) | 0.71 (0.67-0.74) | 0.80 (0.78-0.82) |
| Other backward class | 0.73 (0.71-0.74) | 0.67 (0.65-0.69) | 0.77 (0.76-0.79) | 0.71 (0.68-0.73) | 0.79 (0.78-0.81) |
| MPCE Lowest | 0.75 (0.74-0.77) | 0.55 (0.51-0.59) | 0.85 (0.83-0.87) | 0.62 (0.58-0.66) | 0.79 (0.77-0.81) |
| MPCE Lower middle | 0.73 (0.71-0.75) | 0.58 (0.55-0.62) | 0.82 (0.80-0.84) | 0.67 (0.64-0.70) | 0.79 (0.77-0.81) |
| MPCE Middle | 0.73 (0.71-0.75) | 0.67 (0.64-0.70) | 0.78 (0.75-0.80) | 0.71 (0.68-0.74) | 0.81 (0.79-0.82) |
| MPCE Upper middle | 0.72 (0.70-0.74) | 0.72 (0.69-0.74) | 0.73 (0.70-0.76) | 0.76 (0.74-0.79) | 0.80 (0.78-0.82) |
| MPCE Highest | 0.74 (0.72-0.76) | 0.78 (0.76-0.80) | 0.67 (0.64-0.70) | 0.78 (0.76-0.80) | 0.80 (0.78-0.82) |
| **Fully Convolutional Networks (FCN)** |  |  |  |  |  |
| Overall | 0.69 (0.68-0.69) | 0.67 (0.66-0.69) | 0.70 (0.68-0.71) | 0.65 (0.64-0.67) | 0.76 (0.75-0.76) |
| General | 0.69 (0.67-0.71) | 0.75 (0.73-0.77) | 0.61 (0.58-0.64) | 0.73 (0.71-0.75) | 0.75 (0.73-0.77) |
| Scheduled caste | 0.71 (0.69-0.73) | 0.53 (0.50-0.57) | 0.79 (0.77-0.81) | 0.53 (0.49-0.57) | 0.74 (0.72-0.77) |
| Scheduled tribe | 0.68 (0.66-0.70) | 0.64 (0.60-0.67) | 0.71 (0.68-0.73) | 0.62 (0.59-0.66) | 0.74 (0.71-0.76) |
| Other backward class | 0.67 (0.66-0.69) | 0.67 (0.65-0.69) | 0.68 (0.66-0.70) | 0.63 (0.61-0.65) | 0.74 (0.73-0.76) |
| MPCE Lowest | 0.71 (0.69-0.72) | 0.56 (0.52-0.59) | 0.77 (0.75-0.79) | 0.52 (0.49-0.56) | 0.75 (0.72-0.77) |
| MPCE Lower middle | 0.67 (0.65-0.69) | 0.61 (0.58-0.64) | 0.71 (0.69-0.74) | 0.57 (0.54-0.60) | 0.73 (0.71-0.75) |
| MPCE Middle | 0.68 (0.67-0.70) | 0.67 (0.64-0.70) | 0.70 (0.67-0.72) | 0.64 (0.61-0.67) | 0.75 (0.73-0.77) |
| MPCE Upper middle | 0.69 (0.67-0.71) | 0.70 (0.68-0.73) | 0.67 (0.64-0.70) | 0.72 (0.70-0.75) | 0.75 (0.73-0.77) |
| MPCE Highest | 0.68 (0.66-0.70) | 0.76 (0.73-0.78) | 0.56 (0.53-0.59) | 0.73 (0.70-0.75) | 0.73 (0.71-0.75) |

**Supplementary Table 4.**
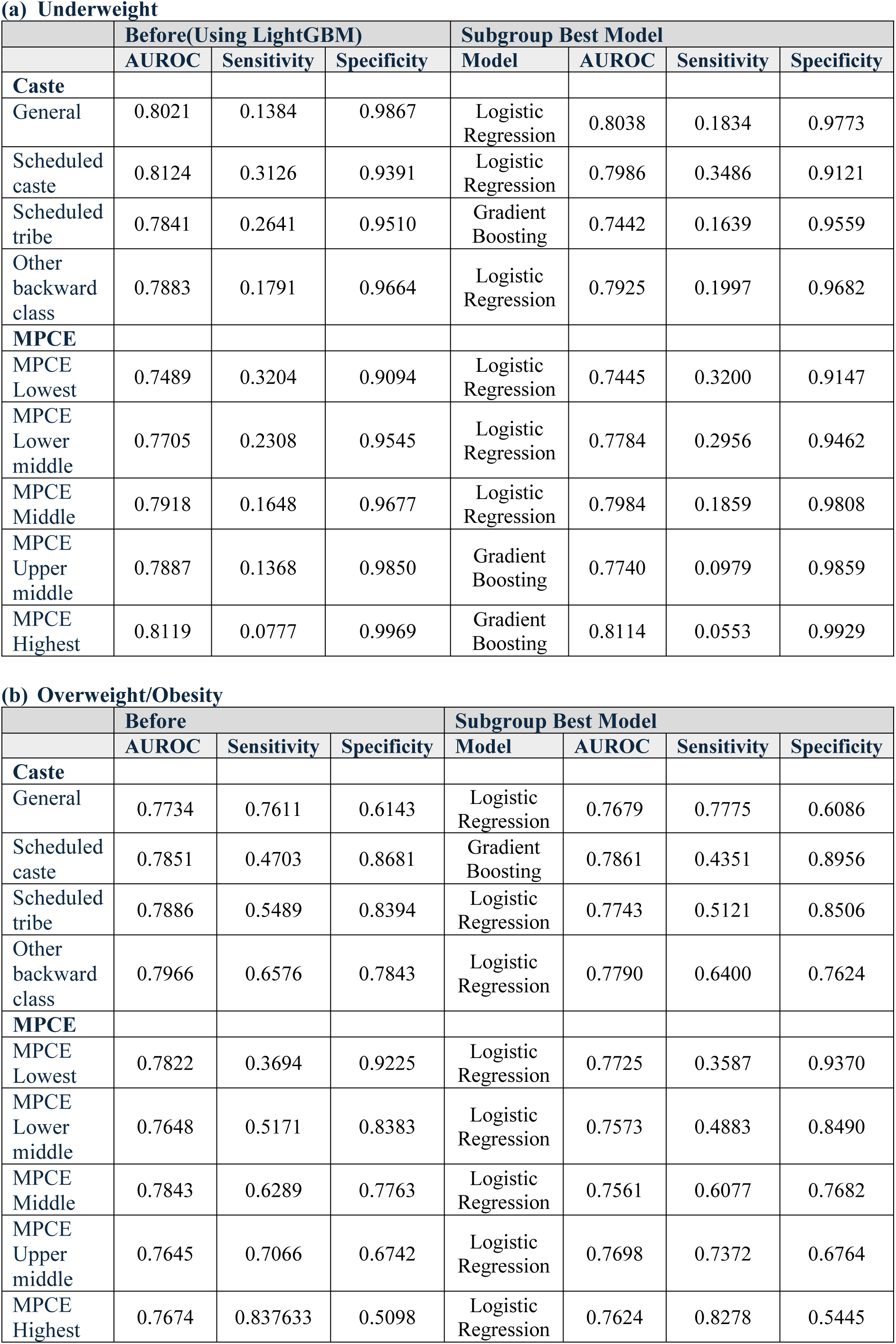
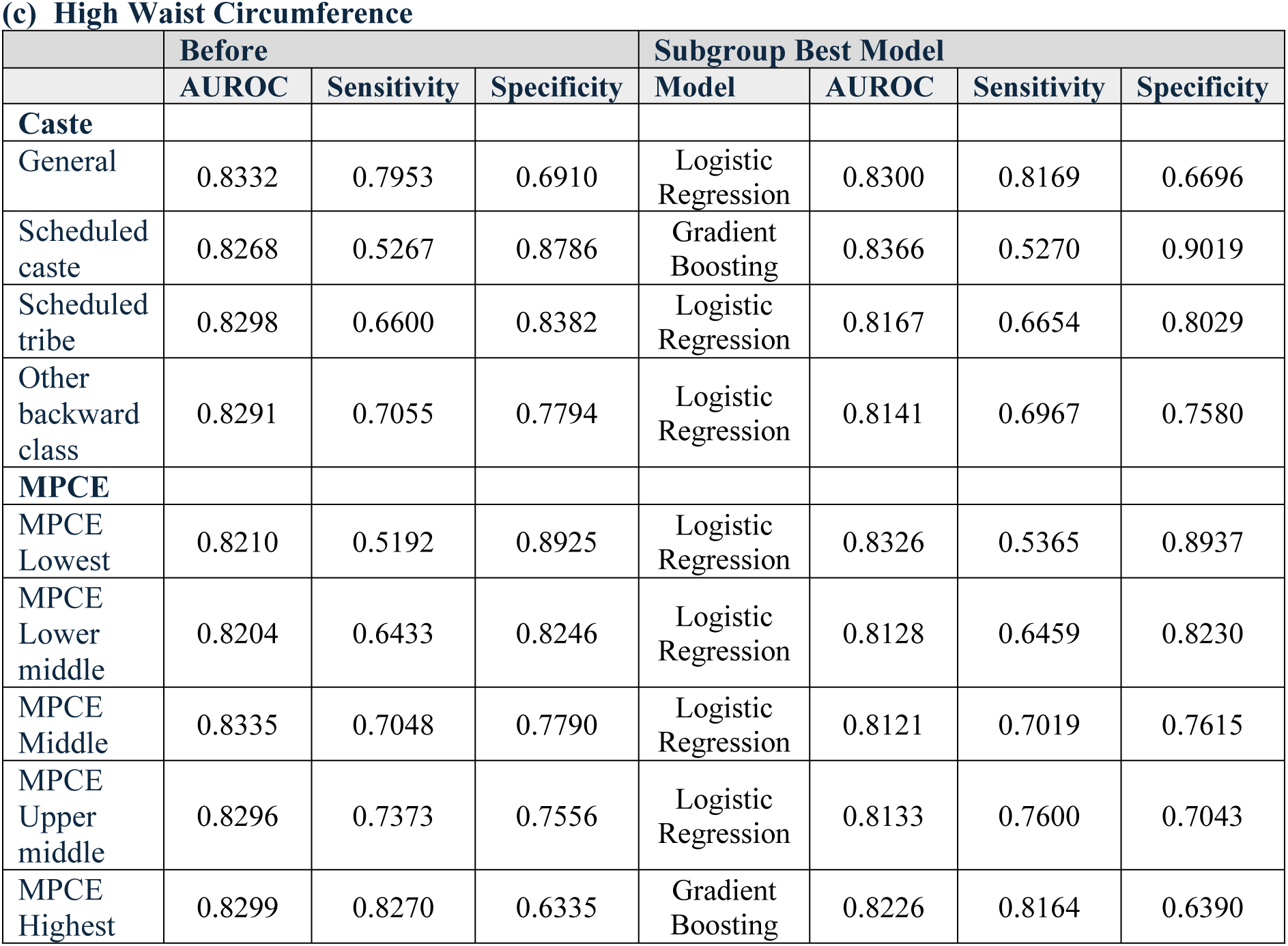
Performance of the Subgroup-Stratified models.

**(a) Underweight**
|  | **Before(Using LightGBM)** |  |  | **Subgroup Best Model** |  |  |  |
| --- | --- | --- | --- | --- | --- | --- | --- |
|  | **AUROC** | **Sensitivity** | **Specificity** | **Model** | **AUROC** | **Sensitivity** | **Specificity** |
| **Caste** |  |  |  |  |  |  |  |
| General | 0.8021 | 0.1384 | 0.9867 | Logistic Regression | 0.8038 | 0.1834 | 0.9773 |
| Scheduled caste | 0.8124 | 0.3126 | 0.9391 | Logistic Regression | 0.7986 | 0.3486 | 0.9121 |
| Scheduled tribe | 0.7841 | 0.2641 | 0.9510 | Gradient Boosting | 0.7442 | 0.1639 | 0.9559 |
| Other backward class | 0.7883 | 0.1791 | 0.9664 | Logistic Regression | 0.7925 | 0.1997 | 0.9682 |
| **MPCE** |  |  |  |  |  |  |  |
| MPCE Lowest | 0.7489 | 0.3204 | 0.9094 | Logistic Regression | 0.7445 | 0.3200 | 0.9147 |
| MPCE Lower middle | 0.7705 | 0.2308 | 0.9545 | Logistic Regression | 0.7784 | 0.2956 | 0.9462 |
| MPCE Middle | 0.7918 | 0.1648 | 0.9677 | Logistic Regression | 0.7984 | 0.1859 | 0.9808 |
| MPCE Upper middle | 0.7887 | 0.1368 | 0.9850 | Gradient Boosting | 0.7740 | 0.0979 | 0.9859 |
| MPCE Highest | 0.8119 | 0.0777 | 0.9969 | Gradient Boosting | 0.8114 | 0.0553 | 0.9929 |

**(b) Overweight/Obesity**
|  | **Before** |  |  | **Subgroup Best Model** |  |  |  |
| --- | --- | --- | --- | --- | --- | --- | --- |
|  | **AUROC** | **Sensitivity** | **Specificity** | **Model** | **AUROC** | **Sensitivity** | **Specificity** |
| **Caste** |  |  |  |  |  |  |  |
| General | 0.7734 | 0.7611 | 0.6143 | Logistic Regression | 0.7679 | 0.7775 | 0.6086 |
| Scheduled caste | 0.7851 | 0.4703 | 0.8681 | Gradient Boosting | 0.7861 | 0.4351 | 0.8956 |
| Scheduled tribe | 0.7886 | 0.5489 | 0.8394 | Logistic Regression | 0.7743 | 0.5121 | 0.8506 |
| Other backward class | 0.7966 | 0.6576 | 0.7843 | Logistic Regression | 0.7790 | 0.6400 | 0.7624 |
| **MPCE** |  |  |  |  |  |  |  |
| MPCE Lowest | 0.7822 | 0.3694 | 0.9225 | Logistic Regression | 0.7725 | 0.3587 | 0.9370 |
| MPCE Lower middle | 0.7648 | 0.5171 | 0.8383 | Logistic Regression | 0.7573 | 0.4883 | 0.8490 |
| MPCE Middle | 0.7843 | 0.6289 | 0.7763 | Logistic Regression | 0.7561 | 0.6077 | 0.7682 |
| MPCE Upper middle | 0.7645 | 0.7066 | 0.6742 | Logistic Regression | 0.7698 | 0.7372 | 0.6764 |
| MPCE Highest | 0.7674 | 0.837633 | 0.5098 | Logistic Regression | 0.7624 | 0.8278 | 0.5445 |

**(c) High Waist Circumference**
|  | **Before** |  |  | **Subgroup Best Model** |  |  |  |
| --- | --- | --- | --- | --- | --- | --- | --- |
|  | **AUROC** | **Sensitivity** | **Specificity** | **Model** | **AUROC** | **Sensitivity** | **Specificity** |
| **Caste** |  |  |  |  |  |  |  |
| General | 0.8332 | 0.7953 | 0.6910 | Logistic Regression | 0.8300 | 0.8169 | 0.6696 |
| Scheduled caste | 0.8268 | 0.5267 | 0.8786 | Gradient Boosting | 0.8366 | 0.5270 | 0.9019 |
| Scheduled tribe | 0.8298 | 0.6600 | 0.8382 | Logistic Regression | 0.8167 | 0.6654 | 0.8029 |
| Other backward class | 0.8291 | 0.7055 | 0.7794 | Logistic Regression | 0.8141 | 0.6967 | 0.7580 |
| **MPCE** |  |  |  |  |  |  |  |
| MPCE Lowest | 0.8210 | 0.5192 | 0.8925 | Logistic Regression | 0.8326 | 0.5365 | 0.8937 |
| MPCE Lower middle | 0.8204 | 0.6433 | 0.8246 | Logistic Regression | 0.8128 | 0.6459 | 0.8230 |
| MPCE Middle | 0.8335 | 0.7048 | 0.7790 | Logistic Regression | 0.8121 | 0.7019 | 0.7615 |
| MPCE Upper middle | 0.8296 | 0.7373 | 0.7556 | Logistic Regression | 0.8133 | 0.7600 | 0.7043 |
| MPCE Highest | 0.8299 | 0.8270 | 0.6335 | Gradient Boosting | 0.8226 | 0.8164 | 0.6390 |

**Supplementary Table 5.** Comparison of Fairness Metrics (Equalized odds) Among Different Bias Mitigation Methods.

|  | (a) Underweight | (b) Overweight/Obesity | (c) High Waist Circumference |
| --- | --- | --- | --- |
| **Basic** |  |  |  |
| Caste | 0.0601 | 0.1382 | 0.1334 |
| MPCE | 0.0863 | 0.2246 | 0.1441 |
| **Disparate Impact Remover** |  |  |  |
| Caste | 0.0601 | 0.1523 | 0.1167 |
| MPCE | 0.0849 | 0.2099 | 0.1370 |
| **Reweighting** |  |  |  |
| Caste | 0.0606 | 0.1419 | 0.1140 |
| MPCE | 0.0790 | 0.1982 | 0.1230 |
| **Prejudice Remover** |  |  |  |
| Caste | 0.0121 | 0.1607 | 0.1741 |
| MPCE | 0.1002 | 0.1023 | 0.0798 |
| **Exponentiated Gradient Reduction** |  |  |  |
| Caste | 0.0310 | 0.0355 | 0.0280 |
| MPCE | 0.0143 | 0.0204 | 0.0326 |
| **Adversarial Debiasing** |  |  |  |
| Caste | 0.0015 | 0.0771 | 0.0167 |
| MPCE | 0.0022 | 0.1278 | 0.0351 |
| **Reject Option Classification** |  |  |  |
| Caste | 0.0424 | 0.0661 | 0.0415 |
| MPCE | 0.0446 | 0.0947 | 0.0476 |
| **Equalized Odds Postprocessing** |  |  |  |
| Caste | 0.0183 | 0.0617 | 0.0515 |
| MPCE | 0.0173 | 0.0701 | 0.0380 |
| **Stratified Subgroup Best Model** |  |  |  |
| Caste | 0.0915 | 0.1696 | 0.1449 |
| MPCE | 0.0955 | 0.2285 | 0.1360 |

